# External validation and updating of clinical severity scores to guide referral of young children with acute respiratory infections in resource-limited primary care settings

**DOI:** 10.1101/2022.12.06.22283016

**Authors:** Arjun Chandna, Lazaro Mwandigha, Constantinos Koshiaris, Direk Limmathurotsakul, Francois Nosten, Yoel Lubell, Rafael Perera-Salazar, Claudia Turner, Paul Turner

## Abstract

**Background:** Accurate and reliable guidelines for referral of children from resource-limited primary care settings are lacking. We identified three practicable paediatric severity scores (Liverpool quick Sequential Organ Failure Assessment [LqSOFA], quick Pediatric Logistic Organ Dysfunction-2 [qPELOD-2], and the modified Systemic Inflammatory Response Syndrome [mSIRS]) and externally validated their performance in young children presenting with acute respiratory infections to a primary care clinic located within a refugee camp on the Thailand-Myanmar border.

**Methods:** This secondary analysis of data from a longitudinal birth cohort study consisted of 3,010 acute respiratory infections in children aged ≤ 24 months. The primary outcome was receipt of supplemental oxygen. We externally validated the discrimination, calibration, and net-benefit of the scores, and quantified gains in performance that might be expected if they were deployed as simple clinical prediction models, and updated to include nutritional status and respiratory distress.

**Results:** 104/3,010 (3.5%) presentations met the primary outcome. The LqSOFA score demonstrated the best discrimination (AUC 0.84; 95% CI 0.79-0.89) and achieved a sensitivity and specificity > 0.80. Converting the scores into clinical prediction models improved performance, resulting in ∼20% fewer unnecessary referrals and ∼30-60% fewer children incorrectly managed in the community.

**Conclusions:** The LqSOFA score is a promising triage tool for young children presenting with acute respiratory infections in resource-limited primary care settings. Where feasible, deploying the score as a simple clinical prediction model might enable more accurate and nuanced risk stratification, increasing applicability across a wider range of contexts.

## INTRODUCTION

Acute respiratory infections (ARIs) are the leading reason for unscheduled childhood medical consultations worldwide.^1,2^ Primary care workers function as gatekeepers to the formal health system, aiming to distinguish the minority of ARIs requiring onward referral from those suitable for community-based care.^3^

In rural regions of many low- and middle-income countries (LMICs) poorly functioning infrastructure, as well as geographic, climatic, socioeconomic, and cultural factors, can complicate referral mechanisms. Particularly in humanitarian and conflict settings referral can entail risks for both patients and providers.^4^ Consequently, there can be substantial inter- and intra-health system variation in referral thresholds.

Existing tools to support community healthcare providers in their assessment of unwell children, such as the World Health Organization’s Integrated Management of Childhood Illnesses (IMCI) and Integrated Community Case Management (iCCM) guidelines, recommend certain ‘Danger Signs’ to guide referrals.^5,6^ However, these lack sensitivity and specificity, and suffer from considerable interobserver variability.^7,8^ A systematic review of paediatric triage tools concluded that none would be reliable in resource-constrained settings and that lack of follow-up data on children managed in the community rendered the validity of existing tools questionable.^9^

In this study we identified paediatric severity scores suitable for use in resource-limited primary care settings and externally validated their ability to guide referral of young children presenting with ARIs.^10^ We characterised the improvement in performance that might be expected if the scores were deployed as simple clinical prediction models and updated to include variables relevant to children presenting with ARIs in rural LMIC settings.

## METHODS

### Study population

Data were collected during a prospective birth cohort study at a medical clinic for refugees and internally displaced people on the Thailand-Myanmar border.^10^ Between September 2007 and September 2008 pregnant women receiving antenatal care at the clinic were invited to participate. Children of consenting women were reviewed at birth and followed-up each month (routine visit) and during any intercurrent illness (illness visit) until 24 months of age. The local circumstances (inability of the population to move freely out of the camp and lack of other medical providers) contributed to low attrition rates and capture of the majority of acute illnesses for which care was sought.

All ARI illness visits were included in this secondary analysis. An ARI was defined as (A) a presentation with rhinorrhoea, nasal congestion, cough, respiratory distress (chest indrawing, nasal flaring, grunting, tracheal tug, and/or head bobbing), stridor, and/or abnormal lung auscultation (crepitations and/or wheeze), and (B) a compatible contemporaneous syndromic diagnosis (rhinitis, croup, bronchiolitis, influenza-like illness, pneumonia, viral infection and/or wheeze) for children sent home directly from the clinic.

### Identification and shortlisting of scores

Drawing on the results of two recent systematic reviews, we longlisted 16 severity scores that might risk stratify young children presenting from the community with acute respiratory infections (Supplementary Table 1).^11,12^ After considering reliability, validity, and feasibility for implementation we excluded eight scores that required specialist equipment and/or laboratory tests unlikely to be practical for the assessment of young children in busy LMIC primary care settings.^13-20^ Four others were excluded as ≥ 25% of the constituent variables were unavailable in the primary dataset.^21-24^ Two of the remaining scores (quick Sequential Organ Failure Assessment [qSOFA] and quick Pediatric Logistic Organ Dysfunction-2 [qPELOD-2]) contained blood pressure.^25,26^ Hypotension is a late sign in paediatric sepsis and not suitable for early recognition of impending serious illness at the community level.^27^ Furthermore, accurate use and maintenance of sphygmomanometers and stethoscopes may not be feasible in resource-limited settings.^28^ Recently, Romaine et al. replaced systolic blood pressure (SBP) with alternate signs of circulatory compromise (heart rate and capillary refill time) to develop the Liverpool-qSOFA (LqSOFA) score, and demonstrated superior performance compared to qSOFA in febrile children presenting from the community.^29^ Hence, we elected to evaluate the LqSOFA score in preference to qSOFA and to evaluate an adapted qPELOD-2 score (replacing SBP with capillary refill time and assessing mental status using the simpler Alert Voice Pain Unresponsive [AVPU] scale rather than the Glasgow Coma Scale [GCS]). The three scores shortlisted for evaluation were the LqSOFA, qPELOD-2, and modified Systemic Inflammatory Response Syndrome (mSIRS) scores (Table 1).^26,29,30^

**TABLE 1.** Shortlisted paediatric severity scores and comparison between original and study populations. bpm = beats or breaths per minute; ED = emergency department; ICU = intensive care unit; PICU = paediatric intensive care unit.

| Score | Constituent variables | Population | Outcome |
| --- | --- | --- | --- |
| **LqSOFA**<sup>29</sup> | 1. Capillary refill time > 2 seconds<br>2. Mental status < alert on AVPU scale<br>3. Heart rate > age-adjusted threshold<br>4. Respiratory rate > age-adjusted threshold<br>*Each variable allocated one point to give score of 0-4* | <u>Derivation</u>: 1,121 febrile children < 16y attending the ED and requiring a blood test at a specialist paediatric hospital in the United Kingdom.<br><u>Validation</u>: 12,241 febrile children < 16y attending the ED at a specialist paediatric hospital in the United Kingdom. | Critical care admission within 48h of ED attendance.<br><u>Prevalence</u>: 4.2% (derivation) and 1.1% (validation). |
| **mSIRS**<sup>30</sup> | 1. Core temperature > 38.5°C or < 36°C<br>2. Heart rate > or < age-adjusted threshold<br>3. Respiratory rate > age-adjusted threshold<br>*Each variable allocated one point to give score of 0-3* | <u>Derivation</u>: expert consensus (original SIRS score).<sup>18</sup><br><u>Validation</u>: 1,184 adults > 18y admitted to a hospital in Sri Lanka with suspected infection. | In-hospital mortality, cardiac arrest or ICU admission (validation).<br><u>Prevalence</u>: 3.6% (validation). |
| **qPELOD-2**<sup>26</sup> | 1. Mental status < 11 on GCS<br>2. Heart rate > age-adjusted threshold<br>3. Blood pressure < age-adjusted threshold<br>*Each variable allocated one point to give score of 0-3* | <u>Derivation</u>: 862 children < 18y admitted to nine European PICUs with suspected infection.<br><u>Validation</u>: 545 children < 18y admitted to a hospital in the Netherlands with suspected bacterial infection.<sup>17</sup> | In-PICU mortality (derivation) or PICU admission and/or mortality (validation).<br><u>Prevalence</u>: 7.0% (derivation) and 3.3% (validation). |
| **This study** | 1. Capillary refill time > 2 seconds<br>2. Mental status < alert on AVPU scale<br>3. Heart rate > age-adjusted threshold<br>4. Respiratory rate > age-adjusted threshold<br>5. Axillary temperature > 38°C or < 35.5°C | 3,010 ARI presentations from 756 children < 2y presenting to a primary care clinic on the Thai-Myanmar border. | Supplemental oxygen therapy.<br><u>Prevalence</u>: 3.5%. |

### Selection of variables for model updating

To update and improve model performance, additional predictors relevant for children presenting with ARIs in LMIC primary care settings were considered for inclusion. Nutritional status (weight-for-age z-score [WAZ]) and presence of respiratory distress were selected a priori, after considering resource constraints, reliability, validity, biological plausibility, availability of data in the primary dataset, and sample size (Supplementary Table 2).^28^

### Data collection

All data were measured by study staff and entered on to structured case report forms. With the exception of anthropometric data, all clinical data were collected at the time of presentation. Core (rectal) temperature was measured for neonates and infants and adjusted to axillary temperature by subtracting 0.5°C.^6^ Mental status was assessed using the AVPU scale. Capillary refill time was measured centrally. For children admitted to the clinic, weight was measured at the time of presentation (seca scale; precision ± 5g for neonates or ± 50g after birth). In addition, all children had their mid-upper arm circumference (MUAC), weight, and height measured at each monthly routine visit. For the purposes of these analyses, age-adjusted z-scores (R package: *z scorer*)^31^ were calculated using the closest anthropometric data to the illness visit within the following window periods: height ≤ 28 days; MUAC ≤ 28 days without intervening admission; weight ≤ 14 days without intervening admission. Median time between the index illness visit and each anthropometric measurement is reported.

### Primary outcome

The primary outcome was receipt of supplemental oxygen during the illness visit. Study staff were unaware which baseline variables were to be used as candidate predictors at the time of ascertaining outcome status. Clinic treatment protocols specified that peripheral oxygen saturation (SpO_2_) must be checked prior to initiation of supplemental oxygen, with therapy only indicated if SpO_2_ was < 90%. All staff were trained on the treatment protocols prior to study commencement.

### Missing data

616 presentations were missing data on one or more candidate predictors (616/3,010; 20.5%) with capillary refill time containing the highest proportion of missingness (442/3,010; 14.7%; Supplementary Table 3). Under a missing-at-random assumption (Supplementary Figure 1), we used multiple imputation with chained equations (MICE) to deal with missing data (R package: *mice*).^32^ Analyses were done in each of 100 imputed datasets and results pooled. Variables included in the imputation model are reported in Supplementary Table 4.

### Statistical methods

We assessed discrimination and calibration of each score by quantifying the area under the receiver operating characteristic curve (AUC) and plotting model scores against observed outcome proportions. We examined predicted classifications at each of the scores’ cut-offs.

Prior to model building we explored the relationship between continuous predictors and the primary outcome using locally-weighted smoothing to identify non-linear patterns. Accordingly, temperature was modelled using restricted cubic splines (R package: *rms*)^33^ with three knots placed at locations based on percentiles (5^th^ and 95^th^) and recognised physiological thresholds (36°C).^34,35^ We used logistic regression to derive the models and tested for important interactions using likelihood ratio tests (LRT). Random-effects were not modelled as 22% (169/756) of children presented only once. All predictors were prespecified and no predictor selection was performed during model development. Internal validation was performed using 100 bootstrap samples with replacement and optimism-adjusted discrimination and calibration reported (R package: *rms*).^33^

Finally, the models were updated by including respiratory distress and WAZ as additional candidate predictors. Penalised (lasso) logistic regression was used for model updating, variable selection, and shrinkage to minimise overfitting (R package: *glmnet*).^36^ A sensitivity analysis confirmed that median imputation grouped by outcome status produced similar results to MICE and hence to avoid conflicts in variable selection across multiply imputed datasets we used this approach to address missing data for model updating (Supplementary Table 5). We assessed discrimination and calibration of the updated models, examined predicted classifications at clinically-relevant referral thresholds, and compared their clinical utility (net-benefit) to the best-performing points-based severity score using decision curve analysis (R package: *dcurves*).^37^ A sensitivity analysis was performed excluding children who were hypoxic at the time of presentation.

All analyses were done in R, version 4.0.2.^38^

### Sample size

No formal sample size calculation for external validation of the existing severity scores was performed. All available data were used to maximise power and generalisability. Of the 3,010 eligible ARI presentations, 104 met the primary outcome, ensuring sufficient outcome events for a robust external validation.^39^ For derivation and updating of the clinical prediction models we followed the methods of Riley et al. and assumed a conservative R^2^ Nagelkerke of 0.15.^40^ At an outcome prevalence of 3.5% (104/3,010) we estimated that up to 13 candidate predictors (events per parameter [EPP] = 8) could be used to build the prediction models whilst minimising the risk of overfitting (R package: *pmsampsize*).^41^

### Ethics and reporting

Ethical approvals were provided by the Mahidol University Ethics Committee (TMEC 21-023) and Oxford Tropical Research Ethics Committee (OxTREC 511-21). The study is reported in accordance with the Transparent Reporting of a multivariable prediction model for Individual Prognosis Or Diagnosis (TRIPOD) guidelines (Supplementary Table 6).^42^

## RESULTS

From September 2007 to September 2008, 999 pregnant women were enrolled, with 965 children born into the cohort. Amongst 4,061 acute illness presentations, 3,064 were for ARIs. Fifty-four ARI presentations were excluded as information on oxygen therapy was not available in the study database, leaving 3,010 presentations from 756 individual children for the primary analysis (Supplementary Figure 2).

Baseline characteristics of the cohort are summarised (Table 2; Supplementary Table 7). The majority of children were managed in the community (72.3%; 2,175/3,010). Median length of stay for the 835 admissions was 3 days (IQR 2 to 4 days). One hundred and four (3.5%; 104/3,010) presentations met the primary outcome, with those with signs of respiratory distress, age-adjusted tachycardia and/or tachypnoea, lower baseline SpO_2_, prolonged capillary refill times, altered mental status, and lower WAZ more likely to require supplemental oxygen (p < 0.001 to 0.014; Table 2).

**TABLE 2.** Baseline characteristics of the cohort stratified by primary outcome status. ^a^Respiratory distress defined as head bobbing, tracheal tug, grunting and/or chest indrawing; ^b^abnormal chest auscultation defined as crepitations and/or wheeze; ^c^rectal temperature converted to axillary temperature for neonates and infants. ^†^Median interval between anthropometric measurement and index illness presentation: length = 8 days (IQR 4-12 days); MUAC = 9 days (IQR 4-13 days); weight = 4 days (IQR 0-10 days). ^*^Missing data: gestation = 5; birthweight = 14; comorbidity = 10; symptom duration = 21; unwell family member = 10; fever = 5; runny nose = 2; noisy breathing = 6; stridor = 1; respiratory distress = 1; head bobbing = 1; tracheal tug = 1; grunting = 1; chest indrawing = 1; abnormal lung auscultation = 59; lung crepitations = 69; wheeze = 79; dehydration = 7; colour = 50; heart rate = 9; respiratory rate = 8; temperature = 3; oxygen saturation = 1,645; capillary refill time = 442; mental status = 37; WLZ = 158; WAZ = 147; MAZ = 682; LAZ = 14.

| Characteristic | Overall<br>N = 3,010<sup>1</sup> | Supplemental oxygen |  | p-value<sup>2</sup> |
| --- | --- | --- | --- | --- |
|  |  | No<br>N = 2,906<sup>1</sup> | Yes<br>N = 104<sup>1</sup> |  |
| Demographics |  |  |  |  |
| Age (months) | 8.1 (3.7, 13.7) | 8.2 (3.8, 13.8) | 7.3 (3.4, 12.7) | 0.40 |
| Male sex | 1,592 / 3,010 (53%) | 1,541 / 2,906 (53%) | 51 / 104 (49%) | 0.40 |
| Birth history |  |  |  |  |
| Gestation (weeks)* | 39.1 (38.1, 40.0) | 39.2 (38.2, 40.0) | 38.4 (37.3, 39.7) | 0.001 |
| Birthweight (kg)* | 2.9 (2.6, 3.2) | 2.9 (2.6, 3.2) | 2.6 (2.0, 3.0) | <0.001 |
| Previous medical history |  |  |  |  |
| Number of previous illness visits | 3.0 (2.0, 6.0) | 3.0 (2.0, 6.0) | 4.0 (2.0, 9.0) | 0.043 |
|  |  | No<br>N = 2,906<sup>1</sup> | Yes<br>N = 104<sup>1</sup> |  |
| Time since last illness visit (days) | 29.0 (3.0, 81.0) | 31.0 (3.0, 82.0) | 11.0 (2.0, 36.5) | <0.001 |
| Number of previous ARI visits | 3.0 (2.0, 5.0) | 3.0 (2.0, 5.0) | 3.5 (2.0, 8.0) | 0.006 |
| Known comorbidity* | 53 / 3,000 (1.8%) | 39 / 2,898 (1.3%) | 14 / 102 (14%) | <0.001 |
| **History of current illness** |  |  |  |  |
| Duration of symptoms (days)* | 3.0 (2.0, 5.0) | 3.0 (2.0, 5.0) | 3.0 (2.0, 5.0) | 0.30 |
| Antibiotics prior to presentation | 145 / 3,010 (4.8%) | 125 / 2,906 (4.3%) | 20 / 104 (19%) | <0.001 |
| Family member unwell* | 287 / 3,000 (9.6%) | 276 / 2,898 (9.5%) | 11 / 102 (11%) | 0.70 |
| **Presenting symptoms and signs** |  |  |  |  |
| Fever* | 1,958 / 3,005 (65%) | 1,885 / 2,901 (65%) | 73 / 104 (70%) | 0.30 |
| Cough | 2,767 / 3,010 (92%) | 2,667 / 2,906 (92%) | 100 / 104 (96%) | 0.11 |
| Runny nose* | 2,565 / 3,008 (85%) | 2,491 / 2,904 (86%) | 74 / 104 (71%) | <0.001 |
| Noisy breathing* | 447 / 3,004 (15%) | 430 / 2,901 (15%) | 17 / 103 (17%) | 0.60 |
| Stridor* | 6 / 3,009 (0.2%) | 6 / 2,905 (0.2%) | 0 / 104 (0%) | >0.90 |
| Respiratory distress<sup>a*</sup> | 508 / 3,009 (17%) | 416 / 2,905 (14%) | 92 / 104 (88%) | <0.001 |
| Head bobbing* | 52 / 3,009 (1.7%) | 27 / 2,905 (0.9%) | 25 / 104 (24%) | <0.001 |
|  |  | No<br>N = 2,906<sup>1</sup> | Yes<br>N = 104<sup>1</sup> |  |
| Tracheal tug<sup>*</sup> | 134 / 3,009 (4.5%) | 96 / 2,905 (3.3%) | 38 / 104 (37%) | <0.001 |
| Grunting<sup>*</sup> | 26 / 3,009 (0.9%) | 11 / 2,905 (0.4%) | 15 / 104 (14%) | <0.001 |
| Chest indrawing<sup>*</sup> | 493 / 3,009 (16%) | 402 / 2,905 (14%) | 91 / 104 (88%) | <0.001 |
| Abnormal lung auscultation<sup>b*</sup> | 1,455 / 2,951 (49%) | 1,372 / 2,852 (48%) | 83 / 99 (84%) | <0.001 |
| Crepitations<sup>*</sup> | 1,158 / 2,941 (39%) | 1,085 / 2,844 (38%) | 73 / 97 (75%) | <0.001 |
| Wheeze<sup>*</sup> | 794 / 2,931 (27%) | 751 / 2,833 (27%) | 43 / 98 (44%) | <0.001 |
| Dehydration<sup>*</sup> | 127 / 3,003 (4.2%) | 121 / 2,899 (4.2%) | 6 / 104 (5.8%) | 0.40 |
| Pale, mottled or cyanosed<sup>*</sup> | 107 / 2,960 (3.6%) | 91 / 2,862 (3.2%) | 16 / 98 (16%) | <0.001 |
| **Vital signs** |  |  |  |  |
| Heart rate (bpm)<sup>*</sup> |  |  |  |  |
| Neonate | 140.0 (132.0, 150.0) | 140.0 (132.0, 148.0) | 150.0 (140.0, 165.0) | 0.014 |
| Infant | 138.0 (128.0, 144.0) | 136.0 (128.0, 144.0) | 147.0 (136.5, 154.0) | <0.001 |
| Child | 128.0 (120.0, 140.0) | 128.0 (120.0, 140.0) | 140.0 (127.5, 149.0) | 0.002 |
| Respiratory rate (bpm)<sup>*</sup> |  |  |  |  |
| Neonate | 48.0 (45.0, 56.0) | 48.0 (44.2, 54.0) | 64.5 (54.0, 77.0) | 0.008 |
|  |  | No<br>N = 2,906<sup>1</sup> | Yes<br>N = 104<sup>1</sup> |  |
| Infant | 48.0 (42.0, 56.0) | 48.0 (42.0, 56.0) | 58.0 (54.0, 66.0) | <0.001 |
| Child | 45.0 (38.0, 52.0) | 44.0 (38.0, 52.0) | 57.0 (46.5, 62.0) | <0.001 |
| Axillary temperature (°C)<sup>c*</sup> | 36.6 (36.0, 37.5) | 36.6 (36.0, 37.4) | 36.8 (36.2, 37.8) | 0.040 |
| Oxygen saturation (%)<sup>*</sup> | 95.0 (93.0, 96.0) | 95.0 (93.0, 96.0) | 88.0 (85.0, 93.0) | <0.001 |
| Capillary refill time > 2 secs<sup>*</sup> | 36 / 2,568 (1.4%) | 27 / 2,476 (1.1%) | 9 / 92 (9.8%) | <0.001 |
| Not alert<sup>*</sup> | 372 / 2,973 (13%) | 306 / 2,875 (11%) | 66 / 98 (67%) | <0.001 |
| **Anthropometrics** |  |  |  |  |
| Weight-for-length z-score (WLZ)<sup>*†</sup> | 0.0 (-0.8, 0.8) | 0.0 (-0.8, 0.8) | -0.5 (-1.8, 0.7) | <0.001 |
| Weight-for-age z-score (WAZ)<sup>*†</sup> | -0.9 (-1.6, -0.2) | -0.9 (-1.6, -0.2) | -1.9 (-3.4, -0.8) | <0.001 |
| MUAC-for-age z-score (MAZ)<sup>*†</sup> | 0.2 (-0.4, 0.8) | 0.2 (-0.4, 0.8) | -0.7 (-1.9, 0.6) | <0.001 |
| Length-for-age z-score (LAZ)<sup>*†</sup> | -1.5 (-2.3, -0.7) | -1.4 (-2.2, -0.7) | -2.4 (-3.4, -1.4) | <0.001 |

<sup>1</sup>Median (IQR); n / N (%)
<sup>2</sup>Wilcoxon rank sum test; Pearson's Chi-squared test; Fisher's exact test

### LqSOFA and qPELOD-2 scores outperform the mSIRS score for risk stratification of ARIs

Discrimination and calibration of the LqSOFA (AUC = 0.84; 95% confidence interval [CI] = 0.79 to 0.89) and qPELOD-2 (AUC = 0.79; 95% CI = 0.74 to 0.84) scores were considerably better than the mSIRS score (AUC = 0.57; 95% CI = 0.51 to 0.63; Figure 1; Supplementary Table 8; Supplementary Figure 3). At a cut-off of ≥ 1 the LqSOFA score demonstrated a sensitivity of 0.80 (95% CI = 0.72 to 0.89) and specificity of 0.86 (95% CI = 0.85 to 0.88); neither the mSIRS nor qPELOD-2 scores achieved a sensitivity and specificity > 0.70 at any cut-off (Table 3).

**TABLE 3.**
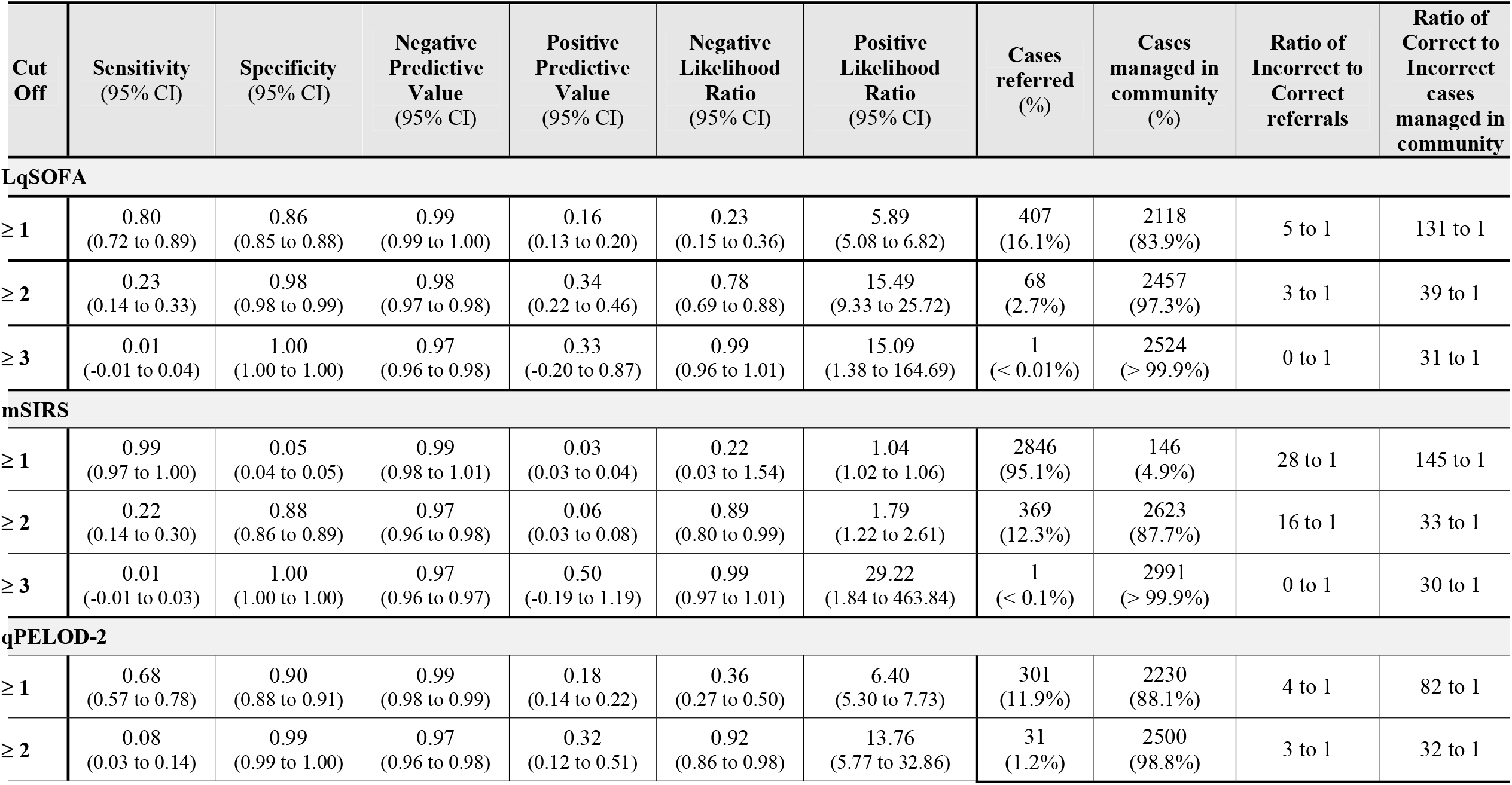
Predicted classifications of the severity scores. Classifications calculated using full-case analysis: LqSOFA = 2,525 presentations (81 met primary outcome); mSIRS = 2,992 presentations (99 met primary outcome); qPELOD-2 = 2,531 presentations (83 met primary outcome)

| **Cut Off** | **Sensitivity**<br>**(95% CI)** | **Specificity**<br>**(95% CI)** | **Negative**<br>**Predictive**<br>**Value**<br>**(95% CI)** | **Positive**<br>**Predictive**<br>**Value**<br>**(95% CI)** | **Negative**<br>**Likelihood**<br>**Ratio**<br>**(95% CI)** | **Positive**<br>**Likelihood**<br>**Ratio**<br>**(95% CI)** | **Cases**<br>**referred**<br>**(%)** | **Cases**<br>**managed in**<br>**community**<br>**(%)** | **Ratio of**<br>**Incorrect to**<br>**Correct**<br>**referrals** | **Ratio of**<br>**Correct to**<br>**Incorrect**<br>**cases**<br>**managed in**<br>**community** |
| --- | --- | --- | --- | --- | --- | --- | --- | --- | --- | --- |
| **LqSOFA** |  |  |  |  |  |  |  |  |  |  |
| **≥ 1** | 0.80<br>(0.72 to 0.89) | 0.86<br>(0.85 to 0.88) | 0.99<br>(0.99 to 1.00) | 0.16<br>(0.13 to 0.20) | 0.23<br>(0.15 to 0.36) | 5.89<br>(5.08 to 6.82) | 407<br>(16.1%) | 2118<br>(83.9%) | 5 to 1 | 131 to 1 |
| **≥ 2** | 0.23<br>(0.14 to 0.33) | 0.98<br>(0.98 to 0.99) | 0.98<br>(0.97 to 0.98) | 0.34<br>(0.22 to 0.46) | 0.78<br>(0.69 to 0.88) | 15.49<br>(9.33 to 25.72) | 68<br>(2.7%) | 2457<br>(97.3%) | 3 to 1 | 39 to 1 |
| **≥ 3** | 0.01<br>(-0.01 to 0.04) | 1.00<br>(1.00 to 1.00) | 0.97<br>(0.96 to 0.98) | 0.33<br>(-0.20 to 0.87) | 0.99<br>(0.96 to 1.01) | 15.09<br>(1.38 to 164.69) | 1<br>(< 0.01%) | 2524<br>(> 99.9%) | 0 to 1 | 31 to 1 |
| **mSIRS** |  |  |  |  |  |  |  |  |  |  |
| **≥ 1** | 0.99<br>(0.97 to 1.00) | 0.05<br>(0.04 to 0.05) | 0.99<br>(0.98 to 1.01) | 0.03<br>(0.03 to 0.04) | 0.22<br>(0.03 to 1.54) | 1.04<br>(1.02 to 1.06) | 2846<br>(95.1%) | 146<br>(4.9%) | 28 to 1 | 145 to 1 |
| **≥ 2** | 0.22<br>(0.14 to 0.30) | 0.88<br>(0.86 to 0.89) | 0.97<br>(0.96 to 0.98) | 0.06<br>(0.03 to 0.08) | 0.89<br>(0.80 to 0.99) | 1.79<br>(1.22 to 2.61) | 369<br>(12.3%) | 2623<br>(87.7%) | 16 to 1 | 33 to 1 |
| **≥ 3** | 0.01<br>(-0.01 to 0.03) | 1.00<br>(1.00 to 1.00) | 0.97<br>(0.96 to 0.97) | 0.50<br>(-0.19 to 1.19) | 0.99<br>(0.97 to 1.01) | 29.22<br>(1.84 to 463.84) | 1<br>(< 0.1%) | 2991<br>(> 99.9%) | 0 to 1 | 30 to 1 |
| **qPELOD-2** |  |  |  |  |  |  |  |  |  |  |
| **≥ 1** | 0.68<br>(0.57 to 0.78) | 0.90<br>(0.88 to 0.91) | 0.99<br>(0.98 to 0.99) | 0.18<br>(0.14 to 0.22) | 0.36<br>(0.27 to 0.50) | 6.40<br>(5.30 to 7.73) | 301<br>(11.9%) | 2230<br>(88.1%) | 4 to 1 | 82 to 1 |
| **≥ 2** | 0.08<br>(0.03 to 0.14) | 0.99<br>(0.99 to 1.00) | 0.97<br>(0.96 to 0.98) | 0.32<br>(0.12 to 0.51) | 0.92<br>(0.86 to 0.98) | 13.76<br>(5.77 to 32.86) | 31<br>(1.2%) | 2500<br>(98.8%) | 3 to 1 | 32 to 1 |

**FIGURE 1.**
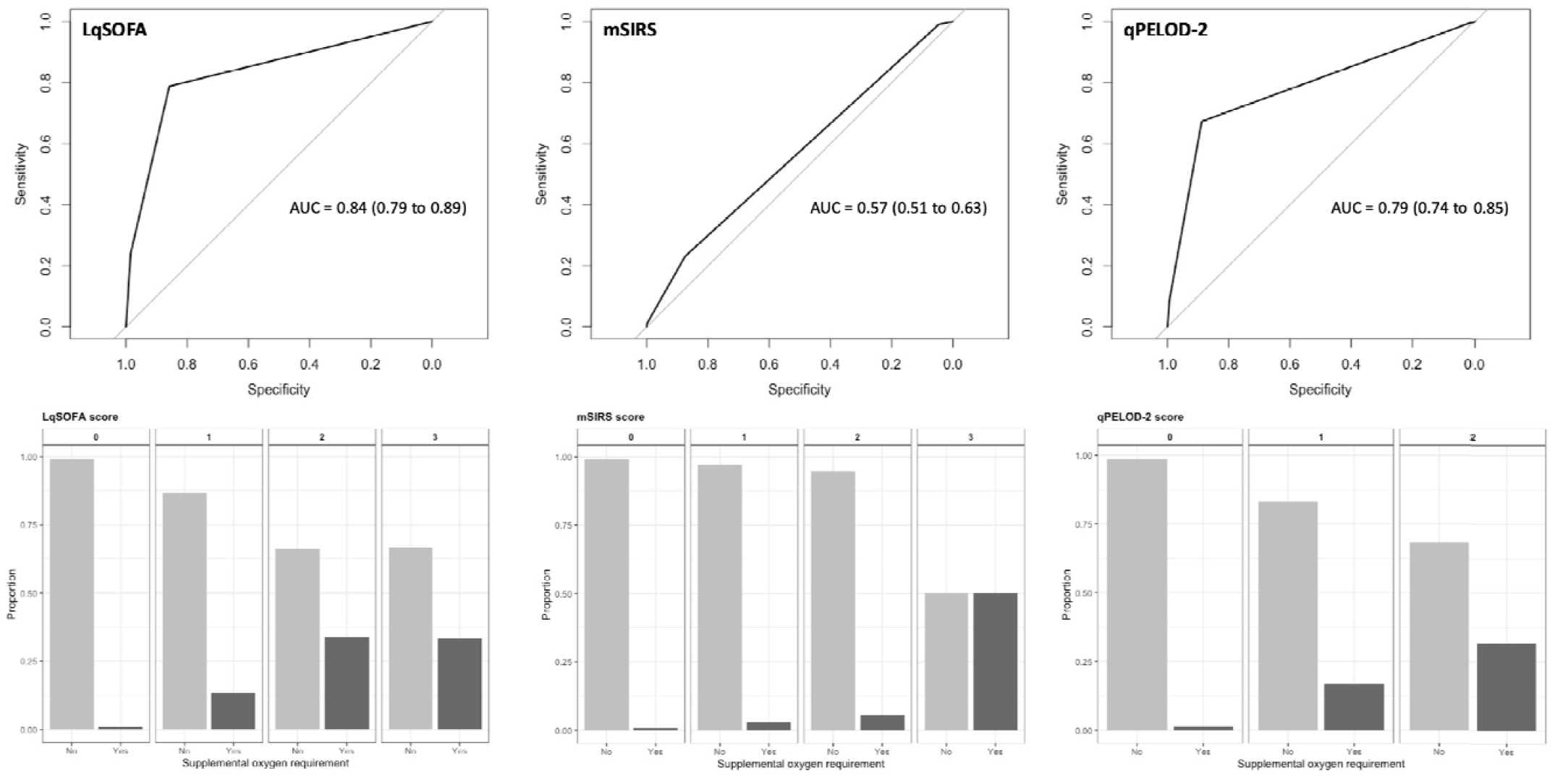
Discrimination of the LqSOFA, mSIRS, and qPELOD-2 severity scores. Receiver operating characteristic curve (ROC) for one imputed dataset shown. Variability in ROCs across multiply imputed datasets shown in Supplementary Figure 2. Pooled AUC reported. Bar plots showing risk scores against observed proportion of oxygen requirement using full case analysis: LqSOFA = 2,525 presentations (81 met primary outcome); mSIRS = 2,992 presentations (99 met primary outcome); qPELOD-2 = 2,531 presentations (83 met primary outcome).

### Improved performance of clinical severity scores when deployed as clinical prediction models

Relationships between continuous predictors and the primary outcome are illustrated (Supplementary Figure 4). There was no evidence of interaction between heart rate (LRT = 2.09; p = 0.35) or respiratory rate (LRT = 0.77; p = 0.68) and age. Optimism-adjusted discrimination of the three models ranged from 0.81 to 0.90, with the LqSOFA model appearing most promising (AUC = 0.90; 95% CI = 0.86 to 0.94; Figure 2; Supplementary Figure 5). Calibration of the qPELOD-2 model was good. The LqSOFA and mSIRS models overestimated risk at higher predicted probabilities.

**FIGURE 2.**
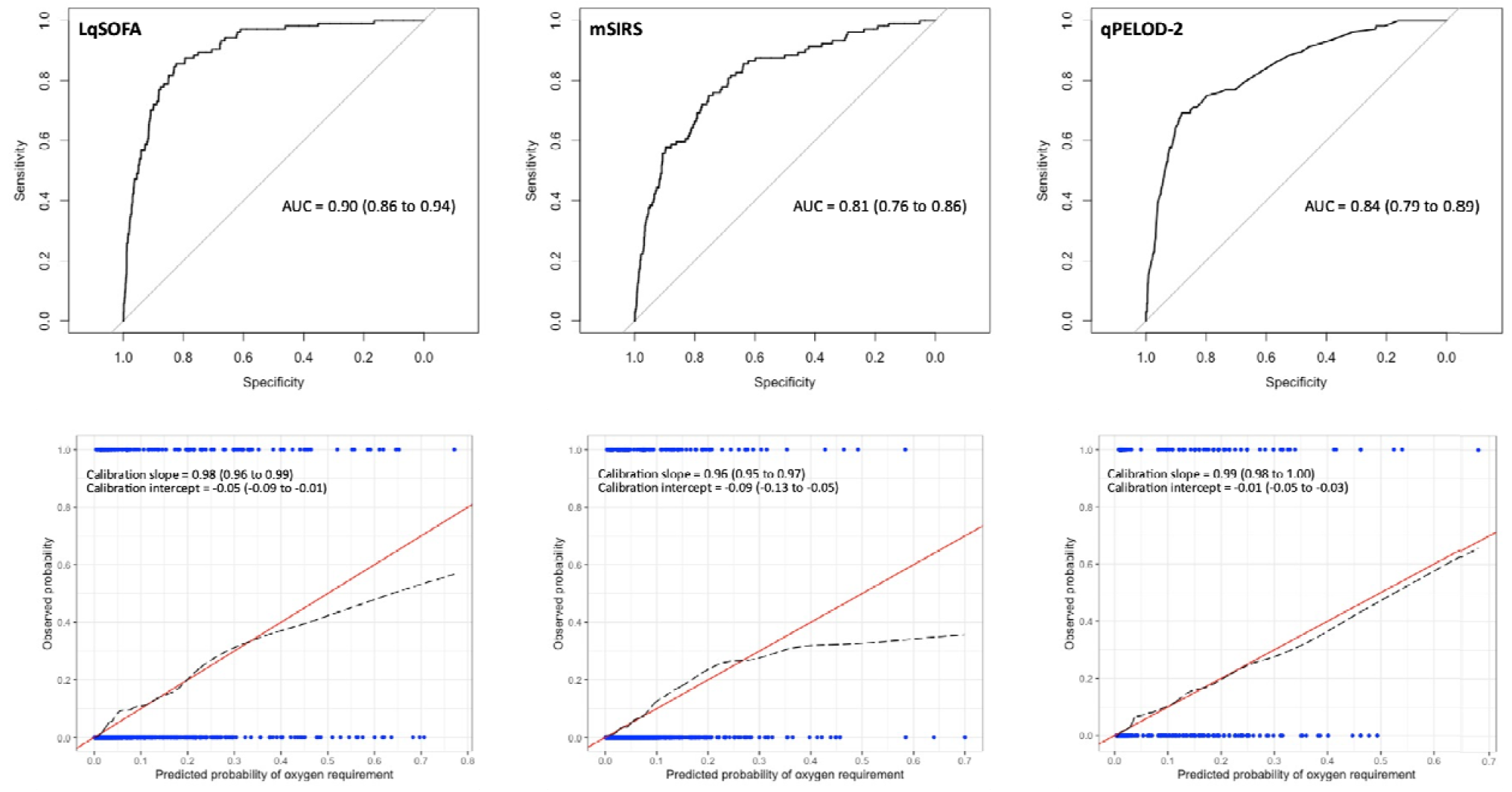
Discrimination and calibration of the LqSOFA, mSIRS, and qPELOD-2 models. Receiver operating characteristic curve (ROC) and calibration slope for one imputed dataset shown. Variability in ROCs and calibration slopes across multiply imputed datasets shown in Supplementary Figure 5. Pooled optimism-adjusted AUCs and calibration slopes reported (100 bootstrap samples). On calibration plots, red line indicates perfect calibration; black dashed line indicates calibration slope for that particular model; blue rug plots indicate distribution of predicted risks for participants who did (top) and did not (bottom) meet the primary outcome.

Discrimination of all three updated models containing respiratory distress and WAZ improved (AUCs = 0.93 to 0.95). Calibration of the updated LqSOFA and qPELOD-2 models was good, whereas the updated mSIRS model underestimated risk at higher predicted probabilities (Figure 3). The full models are reported in Supplementary Table 9.

**FIGURE 3.**
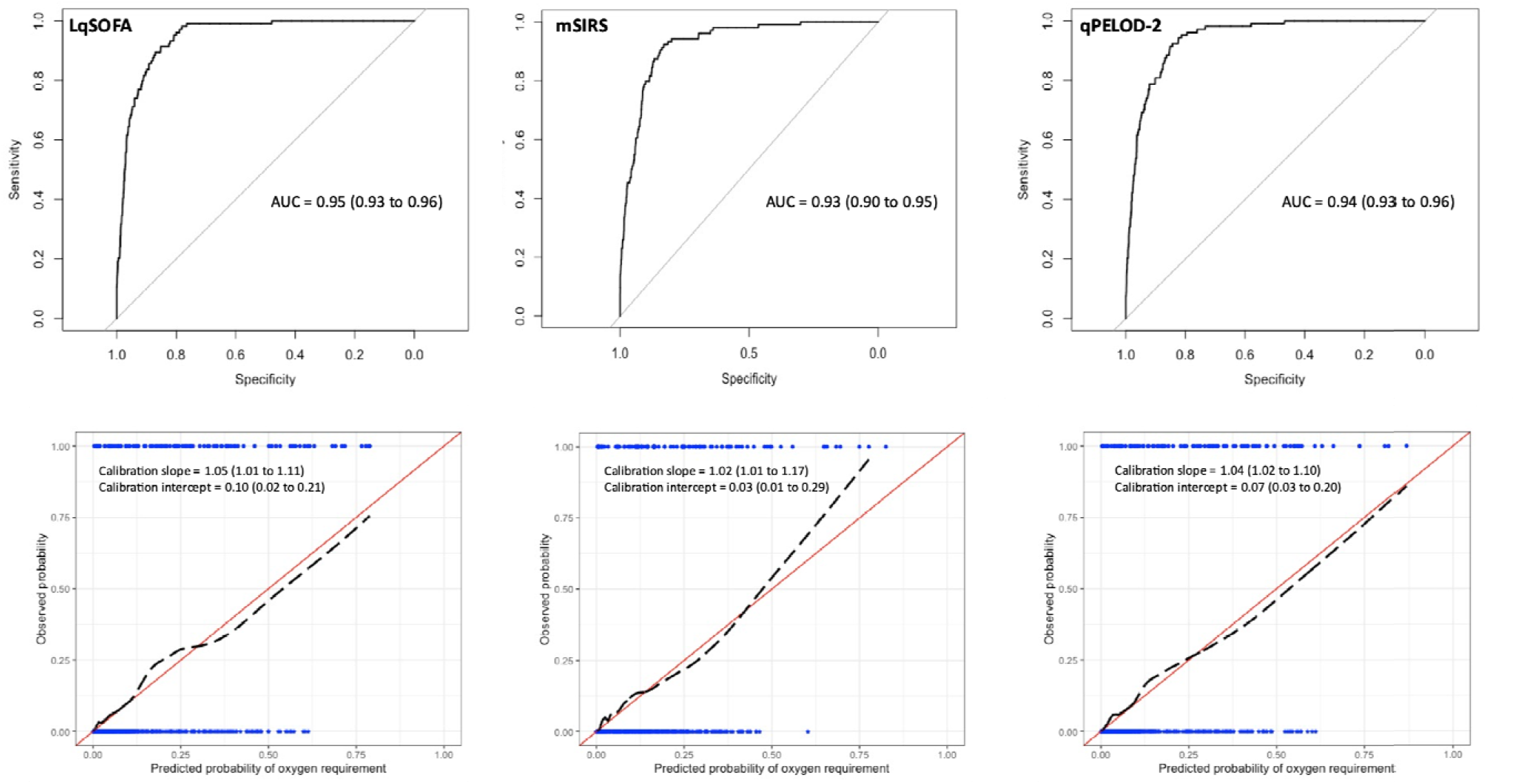
Discrimination and calibration of updated LqSOFA, mSIRS, and qPELOD-2 models. On calibration plots, red line indicates perfect calibration; black dashed line indicates calibration slope for that particular model; blue rug plots indicate distribution of predicted risks for participants who did (top) and did not (bottom) meet the primary outcome.

### Promising clinical utility of the LqSOFA and qPELOD-2 models to guide referrals from primary care

We recognised that the relative value of correct and incorrect referrals is highly context-dependent, reflecting resource availability, practicalities of referral, and capacity for follow-up. Decision curve analyses accounting for differing circumstances suggest that the updated models could provide greater utility (net-benefit) compared to the best points-based score (the LqSOFA score), with the LqSOFA and qPELOD-2 models appearing most promising over a wide range of plausible referral thresholds (Figure 4).

**FIGURE 4.**
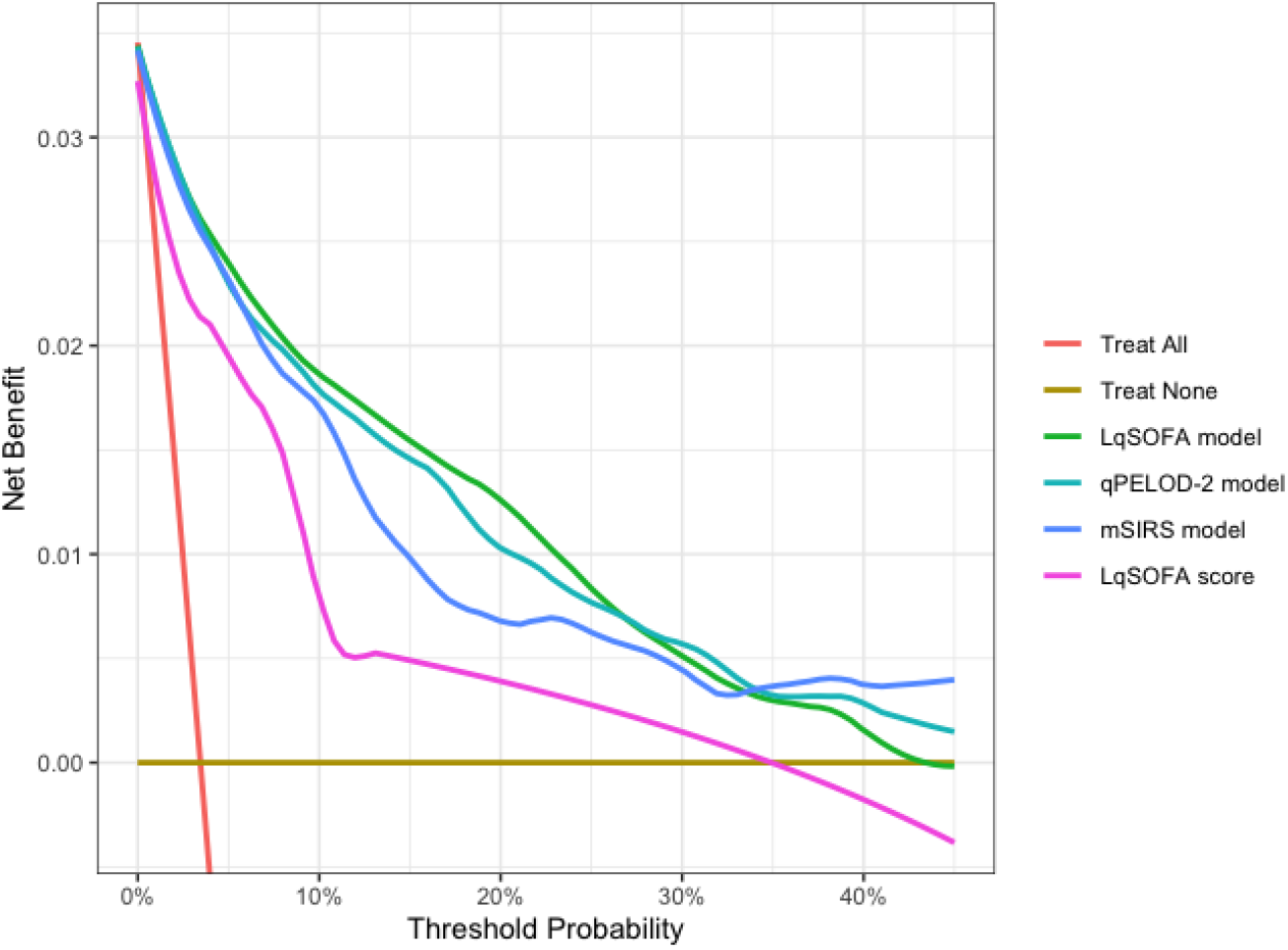
Decision curve analysis of the updated LqSOFA, mSIRS, and qPELOD-2 models. The net benefit of the updated models (green [LqSOFA], turquoise [qPELOD-2], and blue [mSIRS] lines) and original LqSOFA score (pink line), are compared to a “refer-all” (red line) and “refer-none” (brown line) approach. A threshold probability of 5% indicates a management strategy whereby any child with a ≥ 5% probability of requiring oxygen is referred (i.e., a scenario where the value of one correct referral is equivalent to 19 incorrect referrals or a NNR of 20).

The ability of each updated model to guide referrals at thresholds ranging from 1% to 40% is shown (Table 4). A referral threshold of 5% reflects a strategy whereby any child with a predicted probability of requiring oxygen ≥ 5% is referred. At this cut off, the models would suggest referral in ∼15% of all presentations, correctly identifying ∼86-87% of children requiring referral, at a cost of also recommending referral in ∼12-13% of children not requiring referral; i.e., a number needed to refer (NNR; the number of children referred to identify one child who would require oxygen) of five. In contrast, at a similar threshold the LqSOFA score using a cut-off ≥ 1 would suggest referral in a similar proportion of presentations but result in a ∼25% increase in incorrect referrals (a NNR of six) and a ∼30-60% increase in the number of children incorrectly identified for community-based management.

**TABLE 4.** Predicted classifications at different referral thresholds using the updated LqSOFA, qPELOD-2, and mSIRS models. A referral threshold of 5% reflects a management strategy whereby any child with a predicted probability of requiring oxygen ≥ 5% is referred.

| Model | Sensitivity<br>(95% CI) | Specificity<br>(95% CI) | Negative<br>Predictive<br>Value<br>(95% CI) | Positive<br>Predictive<br>Value<br>(95% CI) | Negative<br>Likelihood<br>Ratio<br>(95% CI) | Positive<br>Likelihood<br>Ratio<br>(95% CI) | Cases<br>referred<br>(%) | Cases<br>managed in<br>community<br>(%) | Ratio of<br>Incorrect to<br>Correct<br>referrals | Ratio of<br>Correct to<br>Incorrect cases<br>managed in<br>community |
| --- | --- | --- | --- | --- | --- | --- | --- | --- | --- | --- |
| **Referral threshold = 1%** |  |  |  |  |  |  |  |  |  |  |
| **LqSOFA** | 0.97<br>(0.93 to 1.00) | 0.78<br>(0.73 to 0.82) | 1.00<br>(1.00 to 1.00) | 0.14<br>(0.11 to 0.17) | 0.04<br>(0.00 to 0.09) | 4.48<br>(3.65 to 5.57) | 722<br>(24.0%) | 2288<br>(76.0%) | 6 to 1 | 762 to 1 |
| **qPELOD-2** | 0.96<br>(0.93 to 0.99) | 0.79<br>(0.75 to 0.83) | 1.00<br>(1.00 to 1.00) | 0.14<br>(0.12 to 0.17) | 0.05<br>(0.01 to 0.10) | 4.55<br>(3.97 to 5.87) | 715<br>(23.8%) | 2295<br>(76.2%) | 6 to 1 | 573 to 1 |
| **mSIRS** | 0.94<br>(0.90 to 0.98) | 0.78<br>(0.65 to 0.84) | 1.00<br>(1.00 to 1.00) | 0.13<br>(0.10 to 0.18) | 0.08<br>(0.04 to 0.14) | 4.36<br>(3.00 to 6.21) | 737<br>(24.5%) | 2273<br>(75.5%) | 7 to 1 | 378 to 1 |
| **Referral threshold = 5%** |  |  |  |  |  |  |  |  |  |  |
| **LqSOFA** | 0.87<br>(0.78 to 0.93) | 0.88<br>(0.86 to 0.91) | 0.99<br>(0.99 to 1.00) | 0.21<br>(0.18 to 0.25) | 0.15<br>(0.09 to 0.25) | 7.40<br>(6.22 to 9.50) | 423<br>(14.1%) | 2587<br>(85.9%) | 4 to 1 | 171 to 1 |
| **qPELOD-2** | 0.87<br>(0.78 to 0.93) | 0.87<br>(0.85 to 0.91) | 0.99<br>(0.99 to 1.00) | 0.20<br>(0.16 to 0.23) | 0.15<br>(0.08 to 0.25) | 6.79<br>(5.96 to 8.98) | 468<br>(15.5%) | 2542<br>(84.5%) | 4 to 1 | 181 to 1 |
| **mSIRS** | 0.86<br>(0.77 to 0.93) | 0.87<br>(0.85 to 0.89) | 0.99<br>(0.99 to 1.00) | 0.19<br>(0.16 to 0.22) | 0.16<br>(0.08 to 0.26) | 6.55<br>(5.80 to 7.74) | 470<br>(15.6%) | 2540<br>(84.4%) | 4 to 1 | 211 to 1 |
| **Referral threshold = 10%** |  |  |  |  |  |  |  |  |  |  |
| **LqSOFA** | 0.75<br>(0.66 to 0.83) | 0.93<br>(0.91 to 0.95) | 0.99<br>(0.99 to 0.99) | 0.29<br>(0.24 to 0.36) | 0.26<br>(0.18 to 0.37) | 11.76<br>(9.04 to 16.80) | 270<br>(9.0%) | 2740<br>(91.0%) | 3 to 1 | 100 to 1 |
| **qPELOD-2** | 0.73<br>(0.61 to 0.82) | 0.93<br>(0.90 to 0.95) | 0.99<br>(0.99 to 0.99) | 0.29<br>(0.23 to 0.37) | 0.29<br>(0.20 to 0.41) | 11.57<br>(8.21 to 17.02) | 264<br>(8.8%) | 2764<br>(91.2%) | 3 to 1 | 94 to 1 |
| **mSIRS** | 0.76<br>(0.63 to 0.86) | 0.91<br>(0.88 to 0.93) | 0.99<br>(0.99 to 0.99) | 0.23<br>(0.19 to 0.27) | 0.27<br>(0.16 to 0.41) | 8.22<br>(6.83 to 10.18) | 344<br>(11.4%) | 2666<br>(88.6%) | 3 to 1 | 120 to 1 |
| **Referral threshold = 20%** |  |  |  |  |  |  |  |  |  |  |
| **LqSOFA** | 0.59<br>(0.45 to 0.69) | 0.97<br>(0.96 to 0.97) | 0.99<br>(0.98 to 0.99) | 0.39<br>(0.32 to 0.45) | 0.42<br>(0.32 to 0.56) | 17.82<br>(13.83 to 23.17) | 161<br>(5.3%) | 2849<br>(94.7%) | 2 to 1 | 68 to 1 |
| **qPELOD-2** | 0.56<br>(0.41 to 0.65) | 0.97<br>(0.96 to 0.97) | 0.98<br>(0.98 to 0.99) | 0.37<br>(0.30 to 0.44) | 0.46<br>(0.36 to 0.60) | 16.81<br>(12.98 to 22.87) | 153<br>(5.1%) | 2857<br>(94.9%) | 2 to 1 | 59 to 1 |
| **mSIRS** | 0.49<br>(0.37 to 0.61) | 0.96<br>(0.95 to 0.97) | 0.98<br>(0.98 to 0.99) | 0.31<br>(0.26 to 0.38) | 0.53<br>(0.40 to 0.65) | 12.97<br>(9.85 to 19.44) | 165<br>(5.5%) | 2845<br>(94.5%) | 2 to 1 | 51 to 1 |
| **Referral threshold = 40%** |  |  |  |  |  |  |  |  |  |  |
| **LqSOFA** | 0.28<br>(0.16 to 0.41) | 0.99<br>(0.98 to 1.00) | 0.97<br>(0.97 to 0.98) | 0.49<br>(0.36 to 0.62) | 0.73<br>(0.60 to 0.85) | 27.50<br>(17.38 to 56.63) | 62<br>(2.1%) | 2948<br>(97.9%) | 1 to 1 | 38 to 1 |
| **qPELOD-2** | 0.28<br>(0.13 to 0.41) | 0.99<br>(0.98 to 0.99) | 0.97<br>(0.97 to 0.98) | 0.49<br>(0.35 to 0.59) | 0.73<br>(0.59 to 0.87) | 27.90<br>(16.46 to 47.56) | 62<br>(2.1%) | 2948<br>(97.9%) | 1 to 1 | 39 to 1 |
| **mSIRS** | 0.21<br>(0.09 to 0.35) | 1.00<br>(0.99 to 1.00) | 0.97<br>(0.97 to 0.98) | 0.61<br>(0.47 to 0.90) | 0.80<br>(0.66 to 0.91) | Inf | 20<br>(0.7%) | 2990<br>(99.3%) | 0 to 1 | 35 to 1 |

### Sensitivity analysis

The WHO recommend that pulse oximetry should be universally available at first-level health facilities.^6,43^ Although many barriers exist to realising this laudable goal, to account for the fact that in such contexts a severity score would not be required to guide referral for children who are already hypoxic at the time of presentation, we performed a sensitivity analysis excluding presentations with SpO_2_ < 90% at enrolment. Discrimination remained comparable but clinical utility of the models reduced slightly, with higher NNRs at the lowest referral thresholds (Supplementary Tables 11 & 12).

## DISCUSSION

We report the external validation of three pre-existing severity scores amongst young children presenting with ARIs to a medical clinic on the Thailand-Myanmar border. Unlike other studies which investigated the scores’ prognostic accuracy in hospital settings,^17,25^ we evaluated their performance at the community level and demonstrate that the LqSOFA and qPELOD-2 scores could support early recognition of children requiring referral or closer follow-up in settings with limited resources. In keeping with previous literature, we found that the mSIRS score was poorly discriminative, not well calibrated, and led to substantial misclassification.^17^

An LqSOFA score ≥ 1 yielded a sensitivity and specificity > 80%. Encouragingly, this is remarkably consistent with the performance reported in the original LqSOFA development study and may reflect similarities in the use-case (febrile children presenting from the community) and severity of the cohorts (outcome prevalence 1.1% vs. 3.5%; admission rate 12.1% vs. 27.7%), albeit despite obvious demographic differences.^29^ In contrast to qPELOD-2, LqSOFA contains age-adjusted tachypnoea, which may have improved performance in children with respiratory illnesses. Furthermore, the performance of LqSOFA (or qSOFA) has been shown to improve outside of the PICU, when used to predict more proximal outcomes (e.g. critical care admission rather than mortality), and if the AVPU scale (vs. GCS) is employed to assess mental status.^44^ These all apply to our cohort.

We demonstrated improvement in performance when the severity scores were deployed as clinical prediction models and when nutritional status and respiratory distress were included as additional predictors. Whilst discrimination of all three updated models was good, the AUC is a summary measure of model performance and does not necessarily reflect clinical utility.^45-47^ Decision curve analyses illustrate the superiority of the LqSOFA and qPELOD-2 models compared with the mSIRS model across a range of clinically-relevant referral thresholds.

With growing access to smartphones there may be contexts where the increased accuracy afforded by a clinical prediction model outweighs the simplicity and practicality of points-based scoring systems. At a 5% referral threshold, the updated LqSOFA model identified a similar proportion of presentations for referral as the LqSOFA score at a cut-off of ≥ 1 (14.1% vs. 16.1%), however use of the model would have resulted in ∼25% fewer incorrect referrals and a ∼30% decrease in the number of presentations incorrectly recommended for community-based management. In addition to greater accuracy, prediction models permit more nuanced evaluation of risk; referral thresholds can be adjusted to the needs of an individual patient and/or health system and this flexibility may be particularly impactful in the heterogeneous environments commonplace in many LMIC primary care contexts. For example, in locations where community follow-up is feasible (e.g. via a telephone call or return clinic visit) and/or referral carries great cost (to the patient or system), a higher referral threshold (lower NNR) may be acceptable, compared with settings where safety-netting is impractical and/or access to secondary care is less challenging.

We followed the latest guidelines in prediction model building and used bootstrap internal validation, penalised regression, placed knots at predefined locations, and limited the number of candidate predictors to avoid overfitting the models.^40,42,48,49^ Nevertheless, they require validation on new data to assess generalisability and provide a fairer comparison with the pre-existing points-based scores. We have published our full models to encourage independent validation.

As others have highlighted, a limitation of many studies evaluating community-based triage tools in low-resource settings is the lack of follow-up data for patients categorised as low risk;^9^ 72.3% (2,175/3,010) of our cohort were sent away from the clinic without admission. As acute illness visits were nested within the longitudinal birth cohort, we were able to confirm that 1.4% (30/2,083) of presentations sent away from the clinic without admission received supplemental oxygen within the next 28 days, although it is unknown whether this related to the index ARI or a new illness. A sensitivity analysis conservatively classifying these 30 presentations as meeting the primary outcome (i.e. assuming the oxygen therapy related to the index ARI) resulted in a decrease in the sensitivity of all three models (Supplementary Tables 12 & 13). Prospective research with dedicated outpatient follow-up is ongoing to investigate this issue further.^50^

We selected supplemental oxygen therapy as the primary outcome as this reflects a clinically-meaningful endpoint for ARIs and a pragmatic referral threshold for many resource-limited primary care settings. Oxygen was a scarce resource during the study (cylinders were transported in each week from ∼60km away) and oxygen therapy was protocolised; hence outcome misclassification is less likely.

For those who met the primary outcome, the time of oxygen initiation was not available in the primary dataset. Although no patient had met the outcome when baseline predictors were measured, some may have done so shortly after. Nevertheless, the sensitivity analysis excluding presentations with baseline SpO_2_ < 90% (the qualifying criterion for supplemental oxygen) produced similar results. Furthermore, median length of stay was three days and hence the time horizon for all those who met the primary outcome is likely to have been relatively comparable.

We externally validated three severity scores that could guide assessment of young children presenting with ARIs in resource-limited primary care settings to identify those in need of referral or closer follow-up. Performance of the LqSOFA score was encouraging and comparable to that in the original derivation setting.^29^ Converting the LqSOFA score into a clinical prediction model and including additional variables relevant to resource-constrained LMIC settings improved accuracy and might permit application across a wider range of contexts with differing referral thresholds.

## Data Availability

De-identified, individual participant data from this study will be available to researchers whose proposed purpose of use is approved by the data access committees at the Mahidol-Oxford Tropical Medicine Research Unit. Inquiries or requests for the data may be sent to.

## ABBREVIATIONS

ARI: acute respiratory infection
AUC: area under the receiver operating characteristic curve
AVPU: Alert Voice Pain Unresponsive
CI: confidence interval
EPP: events per parameter
GCS: Glasgow Coma Scale
iCCM: integrated Community Case Management
IMCI: Integrated Management of Childhood Illnesses
IQR: interquartile range
LAZ: length-for-age z-score
LMIC: low- and middle-income country
LqSOFA: Liverpool quick Sequential Organ Failure Assessment
LRT: likelihood ratio test
MAZ: MUAC-for-age z-score
mSIRS: modified Systematic Inflammatory Response Syndrome
MICE: multiple imputation with chained equations
MUAC: mid-upper arm circumference
NNR: number needed to refer
OxTREC: Oxford Tropical Research Ethics Committe
PICU: Paediatric Intensive Care Unit
qPELOD-2: quick Pediatric Logistic Organ Dysfunction-2
qSOFA: quick Sequential Organ Failure Assessment
SBP: systolic blood pressure
SpO_2_: peripheral oxygen saturation
TMEC: Tropical Medicine Ethics Committee
TRIPOD: Transparent Reporting of a multivariable prediction model for Individual Prognosis Or Diagnosis
WAZ: weight-for-age z-score
WLZ: weight-for-length z-score

## CONFLICT OF INTEREST DISCLOSURES

The authors have no conflicts of interest relevant to this article to disclose.

## FUNDING/SUPPORT

This research was funded by the UK Wellcome Trust [219644/Z/19/Z]. RPS acknowledges part support from the NIHR Applied Research Collaboration Oxford & Thames Valley, the NIHR Oxford Medtech and In-Vitro Diagnostics Co-operative and the Oxford Martin School. CK is supported by a Wellcome Trust/Royal Society Sir Henry Dale Fellowship [211182/Z/18/Z]. For the purpose of open access, the author has applied a CC BY public copyright license to any Author Accepted Manuscript version arising from this submission.

## Notes

### Competing Interest Statement

The authors have declared no competing interest.

### Funding Statement

This research was funded by the UK Wellcome Trust [219644/Z/19/Z]. RP acknowledges part support from the NIHR Applied Research Collaboration Oxford & Thames Valley, the NIHR Oxford Medtech and In-Vitro Diagnostics Co-operative and the Oxford Martin School. CK is supported by a Wellcome Trust/Royal Society Sir Henry Dale Fellowship [211182/Z/18/Z]. For the purpose of open access, the author has applied a CC BY public copyright license to any Author Accepted Manuscript version arising from this submission.

### Author Declarations

Ethical approvals were provided by the Mahidol University Ethics Committee (TMEC 21-023) and Oxford Tropical Research Ethics Committee (OxTREC 511-21).

