## Appendix for "External validation and updating of clinical severity scores to guide referral of young children with acute respiratory infections in resource-limited primary care settings"

### SUPPLEMENTARY APPENDIX

**CONTENTS** **PAGE**


| | |
|---|---|
| **1. Longlisted paediatric severity scores** | 2 |
| **2. Candidate variables for updating severity scores** | 3 |
| **3. Proportion of missing data in candidate predictors** | 4 |
| **4. Patterns of missing data in candidate predictors** | 5 |
| **5. Variables included in imputation model** | 6 |
| **6. Comparison of different methods for handling missing data** | 7 |
| **7. TRIPOD checklist** | 8 |
| **8. Eligibility of acute illness visits** | 9 |
| **9. Baseline characteristics for patient-level variables** | 10 |
| **10. Relationship between the severity scores and observed outcome proportions** | 11 |
| **11. Discrimination of the severity scores in each of the multiply imputed datasets** | 13 |
| **12. Relationships between continuous candidate predictors and the primary outcome** | 14 |
| **13. Discrimination and calibration of the models in each of the multiply imputed datasets** | 15 |
| **14. Final model equations for the clinical prediction models** | 16 |
| **15. Sensitivity analysis I: performance of severity scores and models** | 17 |
| **16. Sensitivity analysis I: predicted classifications of updated models** | 18 |
| **17. Sensitivity analysis II: performance of severity scores and models** | 20 |
| **18. Sensitivity analysis II: predicted classifications of updated models** | 21 |

**Supplementary Table 1. Longlisted paediatric severity scores.** Grey shaded rows indicate scores excluded at shortlisting stage. FEAST-PET = Fluid Expansion as Supportive Therapy-Paediatric Emergency Triage; ITAT = Inpatient Triage and Treatment; LODS = Lambarene Organ Dysfunction Score; LqSOFA = Liverpool quick Sequential Organ Failure Assessment; mRISC = modified Respiratory Index of Severity in Children; mSIRS = modified Systemic Inflammatory Response Syndrome; PAWS = Paediatric Advanced Warning Score; PEDIA = Paediatric Early Death Index for Africa; PRISM = Paediatric Risk of Mortality-3; PEWS = Paediatric Early Warning System; qPELOD-2 = quick Pediatric Logistic Organ Dysfunction-2; qSOFA = quick Sequential Organ Failure Assessment; qSOFA-L = qSOFA-Lactate; RISC = Respiratory Index of Severity in Children; SIRS = Systemic Inflammatory Response Syndrome.

| Score | Constituent variables | Included / Excluded |
|---|---|---|
| **FEAST-PET**[1] | Heart rate, temperature, capillary refill time, respiratory distress, lung crepitations, pulse character, prostration, pallor | Excluded – pulse character and prostration not available in primary dataset |
| **ITAT**[2] | Heart rate, temperature, respiratory rate, oxygen saturation | Excluded – pulse oximetry not feasible for young infants in high-throughput LMIC primary care settings |
| **LqSOFA**[3] | Heart rate, respiratory rate, capillary refill time, mental status | Included |
| **LODS**[4] | Deep breathing, coma, prostration | Excluded – deep breathing and prostration not available in primary dataset |
| **mRISC**[5] | Chest indrawing, prostration, weight-for-age z-score, dehydration, history of unconsciousness, night sweats, mental status, lab-confirmed malaria | Excluded – prostration, history of unconsciousness, and night sweats not available in primary dataset. |
| **mSIRS**[6] | Heart rate, respiratory rate, temperature | Included |
| **PAWS**[7] | Heart rate, respiratory rate, temperature, capillary refill time, mental status, oxygen saturation, respiratory distress | Excluded – pulse oximetry not feasible for young infants in high-throughput LMIC primary care settings |
| **PEWS**[8] | Heart rate, respiratory rate, systolic blood pressure, capillary refill time, oxygen saturation, supplemental oxygen, respiratory distress | Excluded – supplemental oxygen therapy not relevant for primary care-based score; pulse oximetry not feasible for young infants in high-throughput LMIC primary care settings |
| **PEDIA(s)**[9] | Anaemia, jaundice, respiratory distress, deep breathing, mental status, prostration, seizures, temperature, wasting, kwashiorkor, symptom duration | Excluded – deep breathing, prostration, and seizures not available in primary dataset |
| **PRISM-III**[10] | Heart rate, temperature, mental status, systolic blood pressure, glucose, potassium, $pCO_2$, pH, acidosis, pupillary reflexes | Excluded – large number of laboratory parameters not appropriate for high-throughput LMIC primary care settings and not available in primary dataset |
| **qPELOD-2**[11] | Heart rate, blood pressure, mental status | Included – blood pressure replaced with capillary refill time; mental status assessed using AVPU instead of GCS |
| **qSOFA**[12] | Respiratory rate, systolic blood pressure, mental status | Excluded – LqSOFA selected instead as capillary refill time and heart rate more appropriate for circulatory assessment in primary care |
| **qSOFA-L**[13] | Respiratory rate, systolic blood pressure, mental status, lactate | Excluded – lactate not feasible for high-throughput LMIC primary care settings and not available in primary database |
| **RISC**[14] | Oxygen saturation, chest indrawing, wheezing, prostration, weight-for-age z-score | Excluded – pulse oximetry not feasible for young infants in high-throughput LMIC primary care settings; prostration not available in primary dataset |
| **RISC-Malawi**[15] | Oxygen saturation, mid-upper arm circumference, sex, wheeze, mental status | Excluded – pulse oximetry not feasible for young infants in high-throughput LMIC primary care settings |
| **SIRS**[16] | Heart rate, respiratory rate, temperature, leukocyte count | Excluded – leukocyte count not feasible for high-throughput LMIC primary care settings; mSIRS selected instead |

**Supplementary Table 2. Candidate variables available in primary dataset considered for updating of existing severity scores.** White shaded rows indicate variables selected for model updating.

| Variables available in primary dataset | Reliability, validity and feasibility considerations for use in LMIC primary care settings |
|---|---|
| **Birthweight** | Validity concerns with regards measurement at time of birth. Reliability concerns with regards recall by caregivers at time of presentation. |
| **Chest auscultation** | Reliability concerns with regards capacity of limited-skill primary care providers for auscultation. Feasibility concerns with regards maintenance of stethoscopes. |
| **Gestational age** | Validity concerns with regards measurement at time of birth. Reliability concerns with regards recall by caregivers at time of presentation. |
| **Nutritional status** | Feasibility and reliability concerns with regards accurate measurement of length (length-for-age and weight-for-length). Validity concerns with regards mid-upper arm circumference. Weight-for-age chosen as best compromise between feasibility, reliability and validity to reflect nutritional status. |
| **Respiratory distress** | Feasibility and validity concerns with regards inclusion of multiple different measures of respiratory distress. Presence of respiratory distress chosen as binary variable for updating of score. |

**Supplementary Table 3. Proportion of missing data in candidate predictors.**

| **Variable** | **Missing data** (N; %) |
| --- | --- |
| Age | 0 |
| Heart rate | 9 (0.3%) |
| Respiratory rate | 8 (0.3%) |
| Axillary temperature | 3 (0.1%) |
| Capillary refill time | 442 (14.7%) |
| Mental status | 37 (0.1%) |
| Weight-for-age z-score | 147 (4.9%) |
| Respiratory distress | 1 (< 0.01%) |

**Supplementary Figure 1. Proportion and pattern of missing data in candidate predictors**. CRT = capillary refill time; WAZ = weight-for-age z-score.

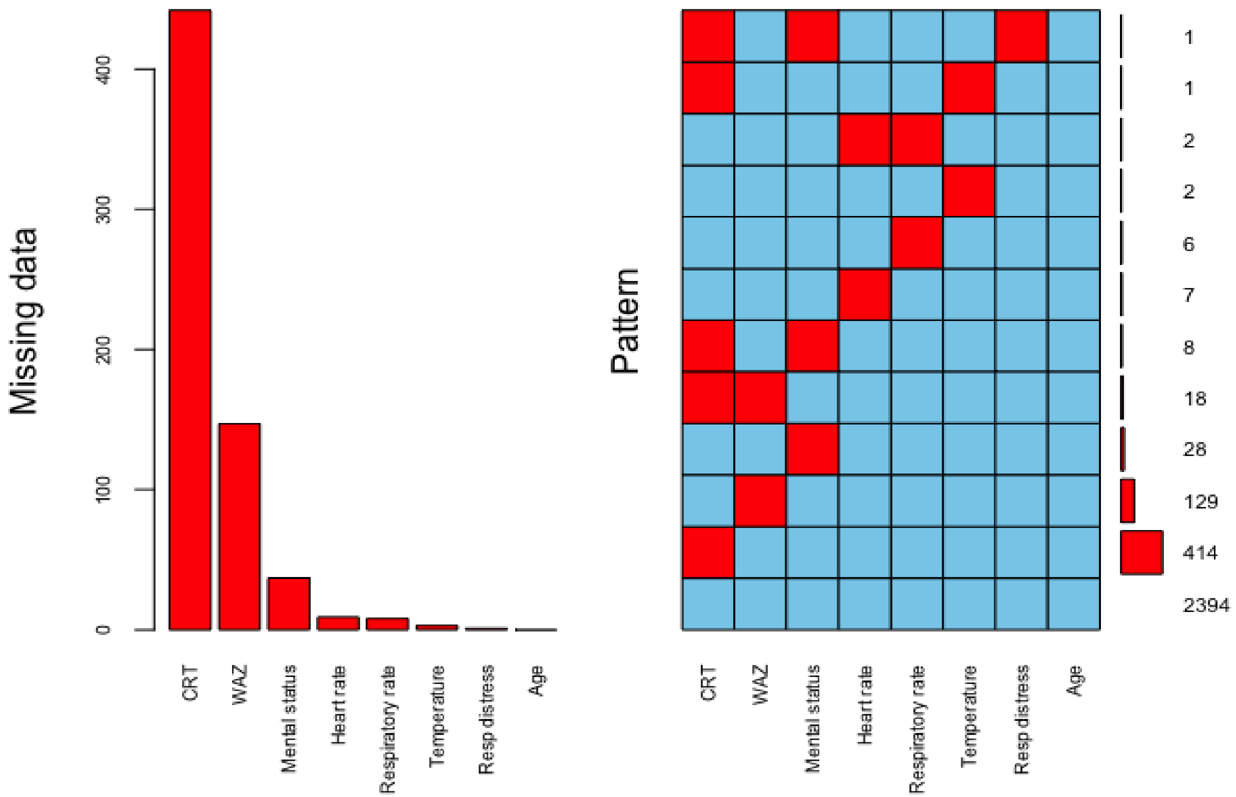

**Supplementary Table 4. Variables included in imputation model.** All explanatory and response variables in the final analysis models, along with variables plausibly associated with capillary refill time (CRT; the variable with the highest proportion of missing data) were included in the imputation model.

| Variable | Reason |
|---|---|
| Age | Final analysis models (explanatory variable) |
| Heart rate | Final analysis models (explanatory variable) |
| Respiratory rate | Final analysis models (explanatory variable) |
| Axillary temperature | Final analysis models (explanatory variable) |
| Capillary refill time | Final analysis models (explanatory variable) |
| Mental status | Final analysis models (explanatory variable) |
| Birthweight | Plausibly associated with CRT |
| Gestational age | Plausibly associated with CRT |
| Known comorbidity | Plausibly associated with CRT |
| Weight-for-age z-score | Final analysis models (explanatory variable) |
| Weight-for-length z-score | Plausibly associated with CRT |
| Length-for-age z-score | Plausibly associated with CRT |
| Head bobbing | Final analysis models (explanatory variable) |
| Nasal flaring | Final analysis models (explanatory variable) |
| Chest indrawing | Final analysis models (explanatory variable) |
| Tracheal tug | Final analysis models (explanatory variable) |
| Grunting | Final analysis models (explanatory variable) |
| Abnormal lung auscultation | Plausibly associated with CRT |
| Dehydration | Plausibly associated with CRT |
| Clinic disposition | Plausibly associated with CRT |
| Receipt of intravenous fluids | Plausibly associated with CRT |
| Receipt of supplemental oxygen | Final analysis models (response variable) |

**Supplementary Table 5. Discrimination of models when missing data addressed using multiple imputation with chained equations, median imputation grouped by outcome status, and a full case analysis.** Dataset for full-case analysis created using pairwise deletion, omitting presentations missing data for any explanatory variable or the response variable (n = 2,523 presentations; 81 outcome events). [*]Pooled discrimination across multiply imputed datasets reported. [†]Optimism-adjusted discrimination reported. MICE = multiple imputation with chained equations.

| Method to addressing missing data | LqSOFA | mSIRS | qPELOD-2 |
|---|---|---|---|
| MICE (95% CI)[*†] | 0.90 (0.86 to 0.94) | 0.81 (0.76 to 0.86) | 0.84 (0.79 to 0.89) |
| Median imputation (95% CI) [†] | 0.89 (0.84 to 0.94) | 0.80 (0.75 to 0.85) | 0.83 (0.78 to 0.88) |
| Full-case analysis (95% CI) [†] | 0.89 (0.84 to 0.94) | 0.80 (0.74 to 0.86) | 0.83 (0.77 to 0.89) |

**Supplementary Table 6. TRIPOD checklist.**

| Section/Topic | | | Checklist Item | Page |
|---|---|---|---|---|
| **Title and abstract** | | | | |
| Title | 1 | D;V | Identify the study as developing and/or validating a multivariable prediction model, the target population, and the outcome to be predicted. | 1 |
| Abstract | 2 | D;V | Provide a summary of objectives, study design, setting, participants, sample size, predictors, outcome, statistical analysis, results, and conclusions. | 3 |
| **Introduction** | | | | |
| Background and objectives | 3a | D;V | Explain the medical context (including whether diagnostic or prognostic) and rationale for developing or validating the multivariable prediction model, including references to existing models. | 4 |
| | 3b | D;V | Specify the objectives, including whether the study describes the development or validation of the model or both. | 4 |
| **Methods** | | | | |
| Source of data | 4a | D;V | Describe the study design or source of data (e.g., randomized trial, cohort, or registry data), separately for the development and validation data sets, if applicable. | 5 |
| | 4b | D;V | Specify the key study dates, including start of accrual; end of accrual; and, if applicable, end of follow-up. | 5 |
| Participants | 5a | D;V | Specify key elements of the study setting (e.g., primary care, secondary care, general population) including number and location of centres. | 5 |
| | 5b | D;V | Describe eligibility criteria for participants. | 5 |
| | 5c | D;V | Give details of treatments received, if relevant. | NA |
| Outcome | 6a | D;V | Clearly define the outcome that is predicted by the prediction model, including how and when assessed. | 7 |
| | 6b | D;V | Report any actions to blind assessment of the outcome to be predicted. | 7 |
| Predictors | 7a | D;V | Clearly define all predictors used in developing or validating the multivariable prediction model, including how and when they were measured. | 5-6 |
| | 7b | D;V | Report any actions to blind assessment of predictors for the outcome and other predictors. | NA |
| Sample size | 8 | D;V | Explain how the study size was arrived at. | 7 |
| Missing data | 9 | D;V | Describe how missing data were handled (e.g., complete-case analysis, single imputation, multiple imputation) with details of any imputation method. | 7 |
| Statistical analysis methods | 10a | D | Describe how predictors were handled in the analyses. | 7-8 |
| | 10b | D | Specify type of model, all model-building procedures (including any predictor selection), and method for internal validation. | 8 |
| | 10c | V | For validation, describe how the predictions were calculated. | 7 |
| | 10d | D;V | Specify all measures used to assess model performance and, if relevant, to compare multiple models. | 7-8 |
| | 10e | V | Describe any model updating (e.g., recalibration) arising from the validation, if done. | 8 |
| Risk groups | 11 | D;V | Provide details on how risk groups were created, if done. | NA |
| Development vs. validation | 12 | V | For validation, identify any differences from the development data in setting, eligibility criteria, outcome, and predictors. | Table 1 |
| **Results** | | | | |
| Participants | 13a | D;V | Describe the flow of participants through the study, including the number of participants with and without the outcome and, if applicable, a summary of the follow-up time. A diagram may be helpful. | 9; S.Fig 1 |
| | 13b | D;V | Describe the characteristics of the participants (basic demographics, clinical features, available predictors), including the number of participants with missing data for predictors and outcome. | 9 |
| | 13c | V | For validation, show a comparison with the development data of the distribution of important variables (demographics, predictors and outcome). | NA |
| Model development | 14a | D | Specify the number of participants and outcome events in each analysis. | 9 |
| | 14b | D | If done, report the unadjusted association between each candidate predictor and outcome. | NA |
| Model specification | 15a | D | Present the full prediction model to allow predictions for individuals (i.e., all regression coefficients, and model intercept or baseline survival at a given time point). | S.Table 8 |
| | 15b | D | Explain how to the use the prediction model. | 10-11 |
| Model performance | 16 | D;V | Report performance measures (with CIs) for the prediction model. | 10 |
| Model-updating | 17 | V | If done, report the results from any model updating (i.e., model specification, model performance). | 10 |
| **Discussion** | | | | |
| Limitations | 18 | D;V | Discuss any limitations of the study (such as nonrepresentative sample, few events per predictor, missing data). | 13-14 |
| Interpretation | 19a | V | For validation, discuss the results with reference to performance in the development data, and any other validation data. | 12 |
| | 19b | D;V | Give an overall interpretation of the results, considering objectives, limitations, results from similar studies, and other relevant evidence. | 12-14 |
| Implications | 20 | D;V | Discuss the potential clinical use of the model and implications for future research. | 13-14 |
| **Other information** | | | | |
| Supplementary information | 21 | D;V | Provide information about the availability of supplementary resources, such as study protocol, Web calculator, and data sets. | 15 |
| Funding | 22 | D;V | Give the source of funding and the role of the funders for the present study. | 15 |

**Supplementary Figure 2. Eligibility of acute illness visits.** ARI = acute respiratory infection.

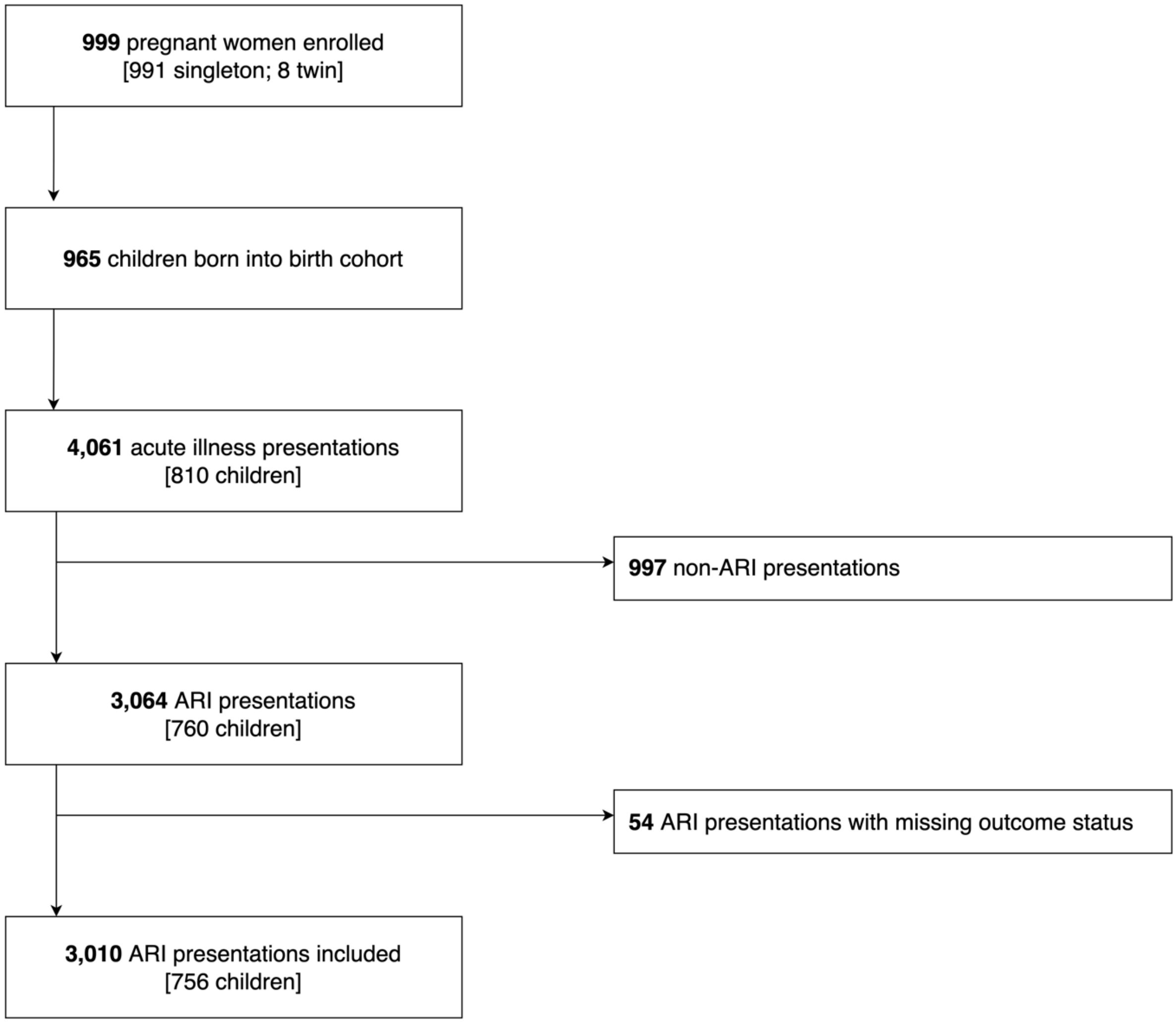

**Supplementary Table 7. Baseline characteristics of the cohort for patient-level variables, stratified by outcome status.** *Missing data: gestation = 2; birthweight = 4.

| Characteristic | Overall N = 756[1] | Supplemental oxygen | | p-value[2] |
|---|---|---|---|---|
| | | No N = 735[1] | Yes N = 21[1] | |
| Male sex | 381 / 756 (50%) | 368 / 735 (50%) | 13 / 21 (62%) | 0.30 |
| Gestation (weeks)* | 39.2 (38.2, 40.0) | 39.2 (38.2, 40.0) | 39.1 (38.0, 40.2) | 0.60 |
| Birthweight (kg)* | 2.9 (2.7, 3.2) | 2.9 (2.7, 3.2) | 2.8 (2.4, 3.2) | 0.40 |

[1]Median (IQR); n / N (%)

[2]Wilcoxon rank sum test; Pearson's Chi-squared test; Fisher's exact test

**Supplementary Table 8. Relationship between the severity scores and observed outcome proportions.** Scores calculated using full-case analysis: LqSOFA = 2,525 presentations; mSIRS = 2,992 presentations; qPELOD-2 = 2,531 presentations. Bpm = beats per minute; LqSOFA = Liverpool quick Sequential Organ Failure Assessment; mSIRS = modified Systemic Inflammatory Response Syndrome; qPELOD-2 = quick Pediatric Logistic Organ Dysfunction-2.

| **Characteristic** | **Overall** N = 3,010[1] | **Supplemental oxygen** | | **p-value**[2] |
|---|---|---|---|---|
| | | **No** N = 2,906[1] | **Yes** N = 104[1] | |
| **LqSOFA** | | | | |
| Respiratory rate > 99th centile[17] | 136 / 3,002 (4.5%) | 111 / 2,901 (3.8%) | 25 / 101 (25%) | <0.001 |
| Heart rate > 99th centile[17] | 7 / 3,001 (0.2%) | 4 / 2,900 (0.1%) | 3 / 101 (3.0%) | 0.001 |
| Not alert | 372 / 2,973 (13%) | 306 / 2,875 (11%) | 66 / 98 (67%) | <0.001 |
| Capillary refill time > 2 secs | 36 / 2,568 (1.4%) | 27 / 2,476 (1.1%) | 9 / 92 (9.8%) | <0.001 |
| ***LqSOFA score*** | | | | |
| 0 | 2,127 / 2,525 (84%) | 2,111 / 2,444 (86%) | 16 / 81 (20%) | <0.001 |
| 1 | 342 / 2,525 (14%) | 296 / 2,444 (12%) | 46 / 81 (57%) | |
| 2 | 53 / 2,525 (2.1%) | 35 / 2,444 (1.4%) | 18 / 81 (22%) | |
| 3 | 3 / 2,525 (0.1%) | 2 / 2,444 (<0.1%) | 1 / 81 (1.2%) | |
| **mSIRS** | | | | |
| Axillary temperature > 38°C | 391 / 3,007 (13%) | 370 / 2,903 (13%) | 21 / 104 (20%) | 0.027 |
| Axillary temperature < 35.5°C | 7 / 3,007 (0.2%) | 5 / 2,903 (0.2%) | 2 / 104 (1.9%) | 0.022 |
| Heart rate > 95th centile | 6 / 3,001 (0.2%) | 3 / 2,900 (0.1%) | 3 / 101 (3.0%) | <0.001 |
| Heart rate < 5th centile | 0 / 3,001 (0%) | 0 / 2,900 (0%) | 0 / 101 (0%) | NA |
| Respiratory rate > 95th centile | 2,852 / 3,002 (95%) | 2,753 / 2,901 (95%) | 99 / 101 (98%) | 0.20 |
| ***mSIRS score*** | | | | |
| 0 | 135 / 2,992 (4.5%) | 134 / 2,893 (4.6%) | 1 / 99 (1.0%) | 0.002 |
| 1 | 2,475 / 2,992 (83%) | 2,399 / 2,893 (83%) | 76 / 99 (77%) | |
| 2 | 380 / 2,992 (13%) | 359 / 2,893 (12%) | 21 / 99 (21%) | |
| 3 | 2 / 2,992 (<0.1%) | 1 / 2,893 (<0.1%) | 1 / 99 (1.0%) | |
| **qPELOD-2** | | | | |
| Heart rate > 195 bpm | 0 / 3,001 (0%) | 0 / 2,900 (0%) | 0 / 101 (0%) | NA |
| Not alert | 372 / 2,973 (13%) | 306 / 2,875 (11%) | 66 / 98 (67%) | <0.001 |
| Capillary refill time > 2 secs | 36 / 2,568 (1.4%) | 27 / 2,476 (1.1%) | 9 / 92 (9.8%) | <0.001 |
| ***qPELOD-2 score*** | | | | |
| 0 | 2,217 / 2,531 (88%) | 2,190 / 2,448 (89%) | 27 / 83 (33%) | <0.001 |

| Characteristic | Overall<br>N = 3,010[1] | Supplemental oxygen | | p-value[2] |
|---|---|---|---|---|
| | | No<br>N = 2,906[1] | Yes<br>N = 104[1] | |
| 1 | 292 / 2,531 (12%) | 243 / 2,448 (9.9%) | 49 / 83 (59%) | |
| 2 | 22 / 2,531 (0.9%) | 15 / 2,448 (0.6%) | 7 / 83 (8.4%) | |

[1]n / N (%)

[2]Fisher's exact test; Pearson's Chi-squared test

**Supplementary Figure 3. Discrimination of the LqSOFA, mSIRS, and qPELOD-2 severity scores in each of the multiply imputed datasets.** Overlay plot to illustrate variability in AUCs across multiply imputed datasets. AUC = area under receiver operating characteristic curve; LqSOFA = Liverpool quick Sequential Organ Failure Assessment; mSIRS = modified Systemic Inflammatory Response Syndrome; qPELOD-2 = quick Pediatric Logistic Organ Dysfunction-2.

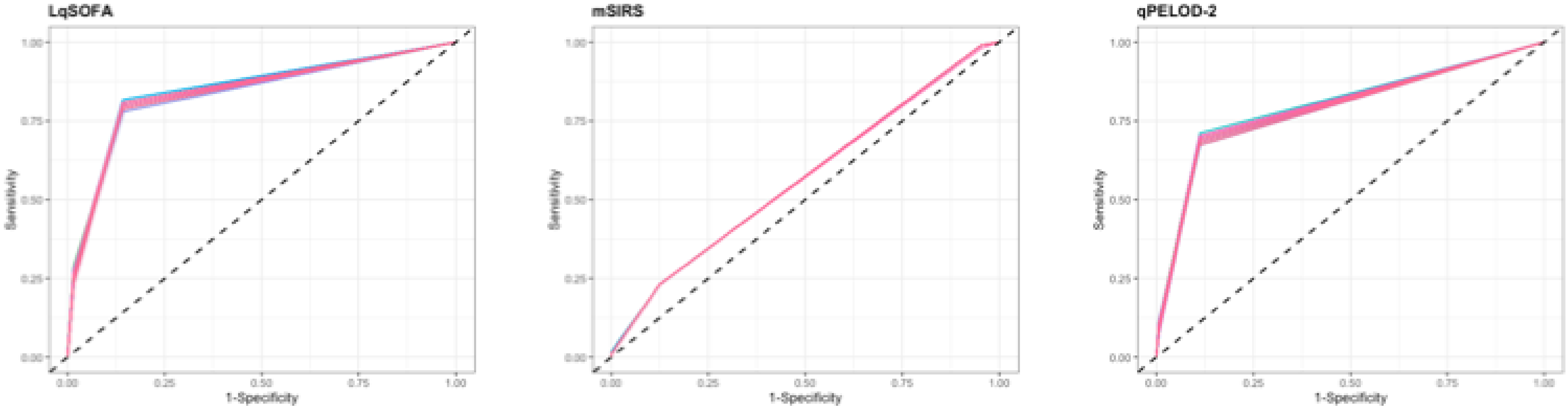

**Supplementary Figure 4. Relationships between continuous candidate predictors and the primary outcome.** Black line = probability of meeting primary outcome; grey shaded area = 95% confidence interval. Bpm = beats / breaths per minute.

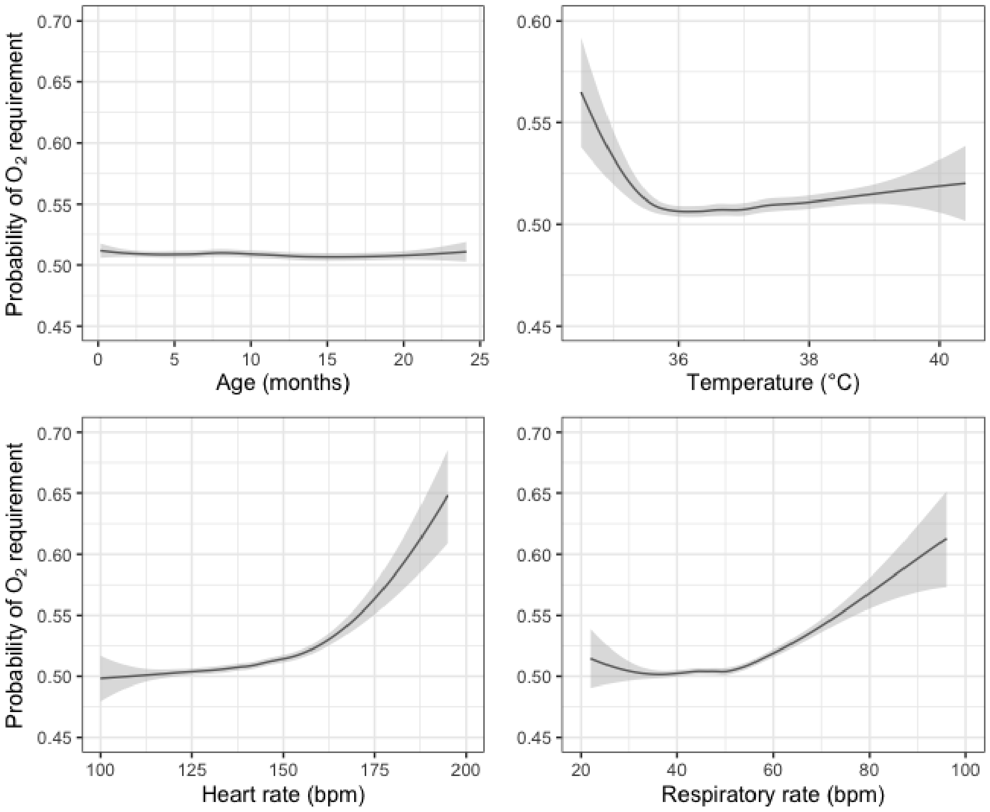

**Supplementary Figure 5. Discrimination and calibration of the LqSOFA, mSIRS, and qPELOD-2 models in each of the multiply imputed datasets.** Overlay plots to illustrate variability in AUCs (top panel) and calibration plots (bottom panel) across multiply imputed datasets. On calibration plots, solid red line indicates perfect calibration; coloured dashed line indicates calibration slope for each imputed dataset; blue rug plots indicate distribution of predicted risks for participants who did (top) and did not (bottom) meet the primary outcome. AUC = area under the receiver operating characteristic curve; LqSOFA = Liverpool quick Sequential Organ Failure Assessment; mSIRS = modified Systemic Inflammatory Response Syndrome; qPELOD-2 = quick Pediatric Logistic Organ Dysfunction-2.

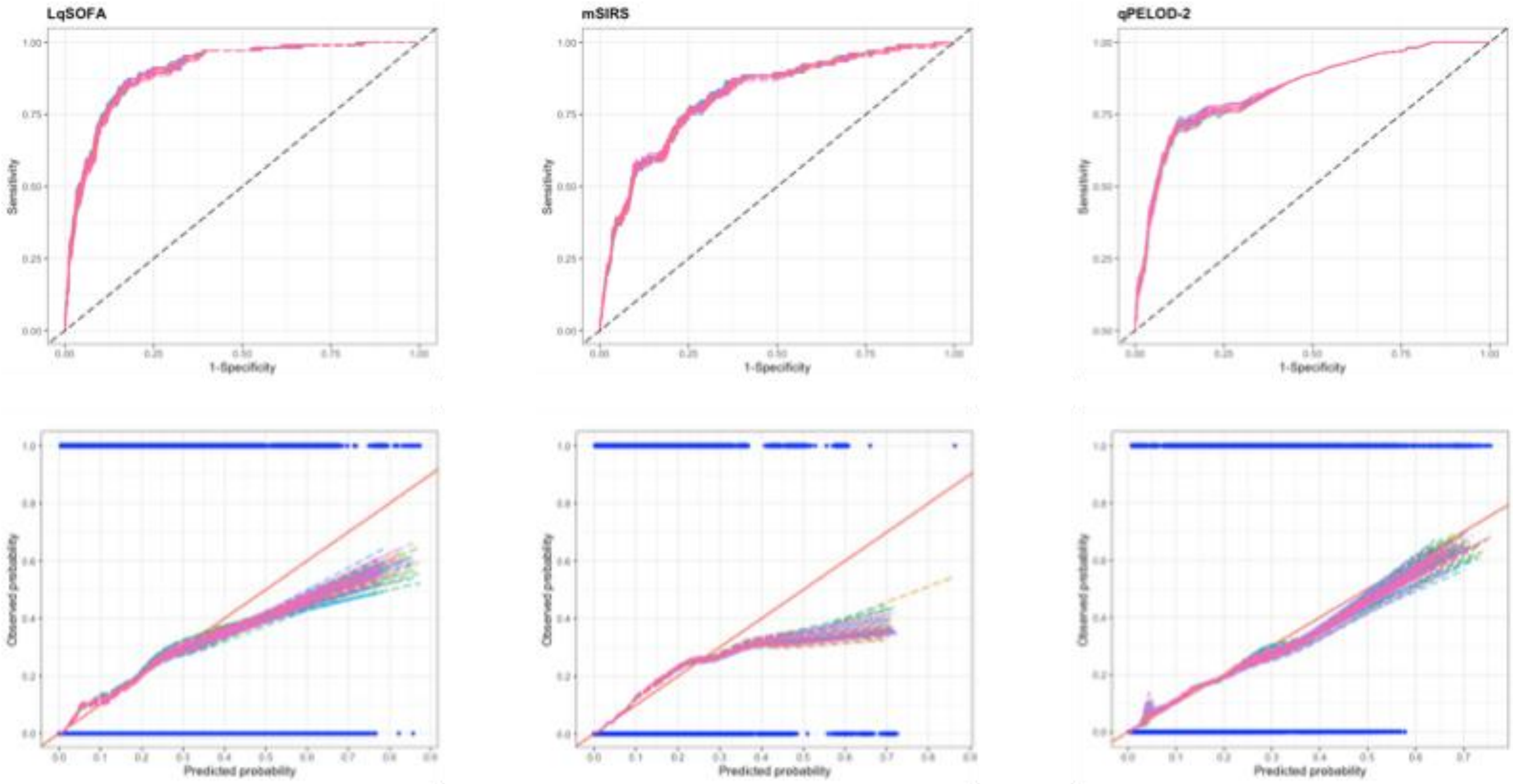

**Supplementary Table 9. Final model equations to illustrate how the clinical prediction models could be used to calculate the predicted probability of supplemental oxygen requirement in children presenting with ARIs in resource-limited primary care settings.**

$$\mathbf{Pr}(\boldsymbol{Oxygen\ requirement}) = \frac{\boldsymbol{e^{LP}}}{\mathbf{1} + \boldsymbol{e^{LP}}}$$

where LP is the linear predictor

The LP should be estimated for each model using the following equations:

<u>**Basic models**</u>

**LqSOFA model**

$$LP(\text{LqSOFA model}) = -11.097 + \begin{cases} 0\ if\ CRT \leq 2 \\ 0.999\ if\ CRT > 2 \end{cases} + \begin{cases} 0\ if\ AVPU = A \\ 2.366\ if\ AVPU < A \end{cases} + 0.025 * age + 0.016 * hr + 0.084 * rr$$

**mSIRS model**

$$LP(\text{mSIRS model}) = 23.819 + \begin{cases} -1.0.47 * temp\ if\ temp < 36 \\ 2.930 * temp\ if\ temp \geq 36 \end{cases} + 0.036 * age + 0.038 * hr + 0.094 * rr$$

**qPELOD-2 model**

$$LP(\text{qPELOD2 model}) = -8.783 + \begin{cases} 0\ if\ CRT \leq 2 \\ 0.966\ if\ CRT > 2 \end{cases} + \begin{cases} 0\ if\ AVPU = A \\ 2.475\ if\ AVPU < A \end{cases} + 0.033 * hr$$

<u>**Updated models**</u>

**LqSOFA model**

$$LP(\text{LqSOFA model}) = -8.727 + \begin{cases} 0\ if\ CRT \leq 2 \\ 0.526\ if\ CRT > 2 \end{cases} + \begin{cases} 0\ if\ AVPU = A \\ 1.685\ if\ AVPU < A \end{cases} - 0.013 * age + 0.008 * hr + 0.035 * rr +$$

$$\begin{cases} 0\ if\ no\ respiratory\ distress \\ 2.722\ if\ respiratory\ distress \end{cases} - 0.364 * WAZ$$

**mSIRS model**

$$LP(\text{mSIRS model}) = 25.623 + \begin{cases} -0.993 * temp\ if\ temp < 36 \\ 3.428 * temp\ if\ temp \geq 36 \end{cases} - 0.024 * age + 0.019 * hr + 0.036 * rr +$$

$$\begin{cases} 0\ if\ no\ respiratory\ distress \\ 3.208\ if\ respiratory\ distress \end{cases} - 0.439 * WAZ$$

**qPELOD-2 model**

$$LP(\text{qPELOD2 model}) = -8.112 + \begin{cases} 0\ if\ CRT \leq 2 \\ 0.558\ if\ CRT > 2 \end{cases} + \begin{cases} 0\ if\ AVPU = A \\ 1.646\ if\ AVPU < A \end{cases} + 0.016 * hr +$$

$$\begin{cases} 0\ if\ no\ respiratory\ distress \\ 3.070\ if\ respiratory\ distress \end{cases} - 0.325 * WAZ$$

<u>Note:</u>
exp is the exponential function

**Supplementary Table 10. Sensitivity analysis I: performance of the severity scores / clinical prediction models when presentations with $SpO_2$ < 90% at enrolment are excluded.** Sensitivity analysis includes 2,949 presentations, 52 of whom met the primary outcome. *Pooled performance measure across multiply imputed datasets reported. †Optimism-adjusted performance measure reported. CI = confidence interval; LqSOFA = Liverpool quick Sequential Organ Failure Assessment; mSIRS = modified Systemic Inflammatory Response Syndrome; qPELOD-2 = quick Pediatric Logistic Organ Dysfunction-2.

| | LqSOFA | mSIRS | qPELOD-2 |
|---|---|---|---|
| **Severity scores (external validation)** | | | |
| Discrimination (95% CI)* | 0.82 (0.74 to 0.89) | 0.55 (0.47 to 0.63) | 0.78 (0.71 to 0.86) |
| **Prognostic models (internal validation)** | | | |
| Discrimination (95% CI)*† | 0.89 (0.83 to 0.95) | 0.78 (0.70 to 0.85) | 0.83 (0.76 to 0.90) |
| Calibration slope (95% CI)*† | 0.95 (0.94 to 0.97) | 0.93 0.91 to 0.95) | 0.98 (0.96 to 1.00) |
| Calibration intercept (95% CI)*† | -0.12 (-0.17 to -0.07) | -0.23 (-0.32 to -0.16) | -0.04 (-0.12 to 0.03) |
| **Prognostic models (updated)** | | | |
| Discrimination (95% CI)† | 0.93 (0.91 to 0.96) | 0.91 (0.87 to 0.93) | 0.93 (0.89 to 0.95) |
| Calibration slope (95% CI)† | 1.08 (1.01 to 1.17) | 1.05 (1.01 to 1.20) | 1.05 (1.01 to 1.17) |
| Calibration intercept (95% CI)† | 0.20 (0.03 to 0.49) | 0.14 (0.02 to 0.61) | 0.14 (0.03 to 0.48) |

**Supplementary Table 11. Sensitivity analysis I: predicted classifications at different referral thresholds using the updated LqSOFA, qPELOD-2, and mSIRS models when presentations with $SpO_2$ < 90% at enrolment are excluded.** Sensitivity analysis includes 2,949 presentations, 52 of whom met the primary outcome. A referral threshold of 5% reflects a management strategy whereby any child with a predicted probability of requiring oxygen ≥ 5% is referred. CI = confidence interval; LqSOFA = Liverpool quick Sequential Organ Failure Assessment; mSIRS = modified Systemic Inflammatory Response Syndrome; qPELOD-2 = quick Pediatric Logistic Organ Dysfunction-2.

| **Model** | **Sensitivity** (95% CI) | **Specificity** (95% CI) | **Negative Predictive Value** (95% CI) | **Positive Predictive Value** (95% CI) | **Negative Likelihood Ratio** (95% CI) | **Positive Likelihood Ratio** (95% CI) | **Cases referred** (%) | **Cases managed in community** (%) | **Ratio of Incorrect to Correct referrals** | **Ratio of Correct to Incorrect cases managed in community** |
|---|---|---|---|---|---|---|---|---|---|---|
| **Referral threshold = 1%** | | | | | | | | | | |
| **LqSOFA** | 0.94 (0.86 to 1.00) | 0.80 (0.76 to 0.85) | 1.00 (1.00 to 1.00) | 0.08 (0.06 to 0.10) | 0.07 (0.02 to 0.17) | 4.68 (3.97 to 6.30) | 628 (21.3%) | 2321 (78.7%) | 12 to 1 | 773 to 1 |
| **qPELOD-2** | 0.94 (0.86 to 0.98) | 0.78 (0.77 to 0.85) | 1.00 (1.00 to 1.00) | 0.08 (0.06 to 0.10) | 0.08 (0.02 to 0.17) | 4.66 (4.05 to 6.53) | 657 (22.3%) | 2292 (77.7%) | 12 to 1 | 763 to 1 |
| **mSIRS** | 0.87 (0.78 to 0.94) | 0.80 (0.70 to 0.85) | 1.00 (1.00 to 1.00) | 0.08 (0.05 to 0.10) | 0.16 (0.07 to 0.29) | 4.60 (3.06 to 6.35) | 596 (20.2%) | 2353 (79.8%) | 12 to 1 | 391 to 1 |
| **Referral threshold = 5%** | | | | | | | | | | |
| **LqSOFA** | 0.69 (0.47 to 0.83) | 0.94 (0.90 to 0.96) | 0.99 (0.99 to 1.00) | 0.17 (0.12 to 0.23) | 0.33 (0.18 to 0.55) | 11.22 (8.04 to 17.80) | 210 (7.1%) | 2739 (92.9%) | 5 to 1 | 170 to 1 |
| **qPELOD-2** | 0.63 (0.44 to 0.79) | 0.94 (0.89 to 0.96) | 0.99 (0.99 to 1.00) | 0.16 (0.11 to 0.23) | 0.39 (0.23 to 0.59) | 10.77 (7.18 to 18.06) | 206 (7.0%) | 2743 (93.0%) | 5 to 1 | 136 to 1 |
| **mSIRS** | 0.71 (0.51 to 0.84) | 0.90 (0.87 to 0.94) | 0.99 (0.99 to 1.00) | 0.12 (0.09 to 0.15) | 0.32 (0.17 to 0.53) | 7.49 (5.94 to 10.93) | 328 (11.1%) | 2621 (88.9%) | 8 to 1 | 186 to 1 |
| **Referral threshold = 10%** | | | | | | | | | | |
| **LqSOFA** | 0.52 (0.34 to 0.68) | 0.96 (0.95 to 0.97) | 0.99 (0.99 to 0.99) | 0.21 (0.15 to 0.27) | 0.50 (0.33 to 0.68) | 14.97 (10.86 to 21.45) | 142 (4.8%) | 2807 (95.2%) | 4 to 1 | 107 to 1 |
| **qPELOD-2** | 0.49 (0.32 to 0.63) | 0.97 (0.96 to 0.97) | 0.99 (0.99 to 0.99) | 0.20 (0.15 to 0.27) | 0.52 (0.38 to 0.70) | 14.39 (10.42 to 21.35) | 112 (3.8%) | 2837 (96.2%) | 3 to 1 | 104 to 1 |
| **mSIRS** | 0.46 (0.25 to 0.64) | 0.96 (0.93 to 0.98) | 0.99 (0.99 to 0.99) | 0.17 (0.12 to 0.22) | 0.56 (0.38 to 0.77) | 11.44 (8.14 to 17.71) | 137 (4.6%) | 2812 (95.4%) | 6 to 1 | 90 to 1 |

| | | | | | | | | | | |
|---|---|---|---|---|---|---|---|---|---|---|
| **Referral threshold = 20%** | | | | | | | | | | |
| **LqSOFA** | 0.31 (0.10 to 0.52) | 0.98 (0.97 to 1.00) | 0.99 (0.98 to 0.99) | 0.27 (0.17 to 0.37) | 0.70 (0.49 to 0.90) | Inf | 76 (2.6%) | 2873 (97.4%) | 3 to 1 | 84 to 1 |
| **qPELOD-2** | 0.32 (0.10 to 0.54) | 0.98 (0.97 to 0.99) | 0.99 (0.98 to 0.99) | 0.28 (0.19 to 0.39) | 0.69 (0.47 to 0.91) | 21.96 (13.93 to 41.68) | 76 (2.6%) | 2873 (97.4%) | 3 to 1 | 84 to 1 |
| **mSIRS** | 0.18 (0.00 to 0.37( | 0.99 (0.98 to 1.00) | 0.99 (0.98 to 0.99) | 0.30 (0.11 to 0.55) | 0.83 (0.64 to 1.00) | Inf | 39 (1.3%) | 2910 (98.7%) | 3 to 1 | 68 to 1 |
| **Referral threshold = 40%** | | | | | | | | | | |
| **LqSOFA** | 0.09 (0.00 to 0.25) | 1.00 (0.99 to 1.00) | 0.98 (0.98 to 0.99) | 0.65 (0.25 to 1.00) | 0.91 (0.76 to 1.00) | Inf | 7 (0.2%) | 2942 (99.8%) | 1 to 1 | 38 to 1 |
| **qPELOD-2** | 0.08 (0.00 to 0.27) | 1.00 (0.99 to 1.00) | 0.98 (0.98 to 0.99) | 0.69 (0.33 to 1.00) | 0.92 (0.74 to 1.00) | Inf | 3 (0.1%) | 2946 (99.9%) | 0 to 1 | 59 to 1 |
| **mSIRS** | 0.04 (0.00 to 0.14) | 1.00 (1.00 to 1.00) | 0.98 (0.98 to 0.99) | 0.70 (0.00 to 1.00) | 0.96 (0.86 to 1.00) | Inf | 3 (0.1%) | 2946 (99.9%) | 0 to 1 | 59 to 1 |

**Supplementary Table 12. Sensitivity analysis II: performance of the severity scores / clinical prediction models when presentations which were sent away from the clinic but required oxygen within the 28 days following enrolment are assumed to have met the primary outcome.** Sensitivity analysis includes 3,010 presentations, 134 of whom met the primary outcome. [*]Pooled performance measure across multiply imputed datasets reported. [†]Optimism-adjusted performance measure reported. CI = confidence interval; LqSOFA = Liverpool quick Sequential Organ Failure Assessment; mSIRS = modified Systemic Inflammatory Response Syndrome; qPELOD-2 = quick Pediatric Logistic Organ Dysfunction-2.

| | **LqSOFA** | **mSIRS** | **qPELOD-2** |
|---|---|---|---|
| **Severity scores (external validation)** | | | |
| Discrimination (95% CI)[*] | 0.75 (0.71 to 0.80) | 0.54 (0.49 to 0.59) | 0.71 (0.66 to 0.76) |
| **Prognostic models (internal validation)** | | | |
| Discrimination (95% CI)[*†] | 0.82 (0.78 to 0.87) | 0.76 (0.71 to 0.81) | 0.77 (0.72 to 0.82) |
| Calibration slope (95% CI)[*†] | 0.98 (0.96 to 0.99) | 0.97 (0.95 to 0.98) | 0.99 (0.98 to 1.00) |
| Calibration intercept (95% CI)[*†] | -0.05 (-0.09 to -0.02) | -0.08 (-0.12 to -0.04) | -0.01 (-0.06 to 0.03) |
| **Prognostic models (updated)** | | | |
| Discrimination (95% CI)[†] | 0.86 (0.82 to 0.89) | 0.85 (0.81 to 0.88) | 0.85 (0.81 to 0.89) |
| Calibration slope (95% CI)[†] | 1.03 (1.01 to 1.09) | 1.02 (1.00 to 1.09) | 1.02 (1.01 to 1.09) |
| Calibration intercept (95% CI)[†] | 0.07 (0.02 to 0.20) | 0.04 (0.01 to 0.22) | 0.05 (0.03 to 0.20) |

**Supplementary Table 13. Sensitivity analysis II: predicted classifications at different referral thresholds using the updated LqSOFA, qPELOD-2, and mSIRS models when presentations which were sent away from the clinic but required oxygen within the 28 days following enrolment are assumed to have met the primary outcome.** Sensitivity analysis includes 3,010 presentations, 134 of whom met the primary outcome. A referral threshold of 5% reflects a management strategy whereby any child with a predicted probability of requiring oxygen ≥ 5% is referred. CI = confidence interval; LqSOFA = Liverpool quick Sequential Organ Failure Assessment; mSIRS = modified Systemic Inflammatory Response Syndrome; qPELOD-2 = quick Pediatric Logistic Organ Dysfunction-2.

| Model | Sensitivity (95% CI) | Specificity (95% CI) | Negative Predictive Value (95% CI) | Positive Predictive Value (95% CI) | Negative Likelihood Ratio (95% CI) | Positive Likelihood Ratio (95% CI) | Cases referred (%) | Cases managed in community (%) | Ratio of Incorrect to Correct referrals | Ratio of Correct to Incorrect cases managed in community |
|---|---|---|---|---|---|---|---|---|---|---|
| **Referral threshold = 1%** | | | | | | | | | | |
| **LqSOFA** | 0.94 (0.89 to 0.98) | 0.36 (0.14 to 0.57) | 0.99 (0.98 to 1.00) | 0.07 (0.05 to 0.09) | 0.17 (0.07 to 0.36) | 1.52 (1.16 to 2.34) | 1931 (64.2%) | 1079 (35.8%) | 14 to 1 | 153 to 1 |
| **qPELOD-2** | 0.95 (0.86 to 0.98) | 0.30 (0.08 to 0.63) | 0.99 (0.97 to 1.00) | 0.06 (0.05 to 0.10) | 0.19 (0.09 to 0.63) | 1.41 (1.08 to 2.72) | 2191 (72.3%) | 819 (27.2%) | 16 to 1 | 163 to 1 |
| **mSIRS** | 0.95 (0.90 to 0.99) | 0.34 (0.14 to 0.52) | 0.99 (0.98 to 1.00) | 0.06 (0.05 to 0.08) | 0.16 (0.06 to 0.33) | 1.46 (1.14 to 2.09) | 2054 (68.2%) | 956 (31.8%) | 15 to 1 | 136 to 1 |
| **Referral threshold = 5%** | | | | | | | | | | |
| **LqSOFA** | 0.74 (0.68 to 0.81) | 0.84 (0.82 to 0.89) | 0.99 (0.98 to 0.99) | 0.19 (0.15 to 0.21) | 0.31 (0.23 to 0.39) | 4.69 (3.92 to 5.71) | 569 (18.9%) | 2441 (81.1%) | 5 to 1 | 65 to 1 |
| **qPELOD-2** | 0.75 (0.68 to 0.81) | 0.83 (0.81 to 0.86) | 0.99 (0.98 to 0.99) | 0.17 (0.15 to 0.21) | 0.30 (0.22 to 0.38) | 4.56 (3.77 to 5.53) | 582 (19.3%) | 2428 (80.7%) | 5 to 1 | 70 to 1 |
| **mSIRS** | 0.75 (0.68 to 0.80) | 0.84 (0.82 to 0.86) | 0.99 (0.98 to 0.99) | 0.18 (0.16 to 0.22) | 0.30 (0.23 to 0.39) | 4.83 (4.10 to 5.64) | 529 (17.6%) | 2481 (82.4%) | 4 to 1 | 68 to 1 |
| **Referral threshold = 10%** | | | | | | | | | | |
| **LqSOFA** | 0.65 (0.55 to 0.73) | 0.91 (0.88 to 0.93) | 0.98 (0.98 to 0.99) | 0.25 (0.21 to 0.29) | 0.38 (0.29 to 0.49) | 7.34 (6.00 to 9.32) | 349 (11.6%) | 2661 (88.4%) | 3 to 1 | 56 to 1 |
| **qPELOD-2** | 0.65 (0.55 to 0.74) | 0.91 (0.87 to 0.93) | 0.98 (0.98 to 0.99) | 0.24 (0.20 to 0.29) | 0.39 (0.29 to 0.49) | 7.02 (5.45 to 9.36) | 354 (11.8%) | 2656 (88.2%) | 3 to 1 | 54 to 1 |
| **mSIRS** | 0.64 (0.52 to 0.73) | 0.89 (0.87 to 0.92) | 0.98 (0.98 to 0.99) | 0.22 (0.19 to 0.26) | 0.40 (0.30 to 0.52) | 6.11 (5.03 to 7.42) | 402 (13.4%) | 2608 (86.6%) | 4 to 1 | 53 to 1 |

| | | | | | | | | | | |
|---|---|---|---|---|---|---|---|---|---|---|
| **Referral threshold = 20%** | | | | | | | | | | |
| **LqSOFA** | 0.48 (0.35 to 0.59) | 0.96 (0.95 to 0.97) | 0.98 (0.97 to 0.98) | 0.37 (0.31 to 0.44) | 0.54 (0.43 to 0.67) | 13.01 (9.55 to 16.90) | 174 (5.8%) | 2836 (94.2%) | 2 to 1 | 39 to 1 |
| **qPELOD-2** | 0.46 (0.33 to 0.55) | 0.96 (0.95 to 0.97) | 0.97 (0.97 to 0.98) | 0.37 (0.30 to 0.44) | 0.56 (0.46 to 0.69) | 13.00 (9.93 to 17.94) | 162 (5.4%) | 2848 (94.6%) | 2 to 1 | 38 to 1 |
| **mSIRS** | 0.41 (0.30 to 0.53) | 0.96 (0.94 to 0.97) | 0.96 (0.94 to 0.97) | 0.30 (0.26 to 0.37) | 0.62 (0.49 to 0.73) | 9.71 (7.42 to 13.84) | 167 (5.5%) | 2843 (94.5%) | 2 to 1 | 34 to 1 |
| **Referral threshold = 40%** | | | | | | | | | | |
| **LqSOFA** | 0.19 (0.10 to 0.29) | 0.99 (0.99 to 1.00) | 0.96 (0.96 to 0.97) | 0.53 (0.40 to 0.68) | 0.82 (0.71 to 0.90) | Inf | 49 (1.6%) | 2961 (98.4%) | 1 to 1 | 25 to 1 |
| **qPELOD-2** | 0.18 (0.08 to 0.28) | 0.99 (0.99 to 1.00) | 0.96 (0.96 to 0.97) | 0.52 (0.40 to 0.68) | 0.82 (0.73 to 0.93) | Inf | 46 (1.5%) | 2964 (98.5%) | 1 to 1 | 25 to 1 |
| **mSIRS** | 0.15 (0.06 to 0.26) | 1.00 (0.99 to 1.00) | 0.96 (0.96 to 0.97) | 0.65 (0.48 too 0.98) | 0.85 (0.74 to 0.94) | Inf | 20 (0.7%) | 2990 (99.3%) | 0 to 1 | 25 to 1 |
